# Recovery-associated resting-state activity and connectivity alterations in Anorexia nervosa

**DOI:** 10.1101/2020.06.21.20135566

**Authors:** Leon D. Lotter, Georg von Polier, Jan Offermann, Kimberly Buettgen, Lukas Stanetzky, Simon B. Eickhoff, Kerstin Konrad, Jochen Seitz, Juergen Dukart

## Abstract

**Background:** Previous studies provided controversial insight on the impact of starvation, disease status and underlying grey matter volume (GMV) changes on resting-state functional magnetic resonance imaging (rsfMRI) alterations in Anorexia nervosa (AN). Here we adapt a combined longitudinal and cross-sectional approach to disentangle the effects of these factors on resting-state alterations in AN.

**Methods:** Overall, 87 female subjects were included in the study: adolescent patients with acute AN scanned at inpatient admission (*N* = 22, mean age 15.3 years) and at discharge (*N* = 21), 21 patients recovered from AN (22.3 years) and two groups of healthy age-matched controls (both *N* = 22, 16.0 and 22.5 years). Whole-brain measures of resting-state activity and functional connectivity were computed (Network Based Statistics, Global Correlation, Integrated Local Correlation, fractional Amplitude of Low Frequency Fluctuations) to assess rsfMRI alterations over the course of AN treatment before and after controlling for underlying GMV.

**Results:** Patients with acute AN displayed strong and widespread prefrontal, sensorimotor, parietal, temporal, precuneal and insular reductions of resting-state connectivity and activity. All alterations were independent of GMV and were largely normalized in short- and absent in long-term recovered AN.

**Conclusions:** Resting-state fMRI alterations in AN constitute acute and GMV independent presumably starvation-related phenomena. The majority of alterations found here normalized over the course of recovery without evidence for possible preexisting trait- or remaining “scar”-effects.

## Introduction

Anorexia nervosa (AN) is a serious eating disorder, typically occurring in females during puberty. It is characterized by a self-induced restriction of food intake and an intense fear of gaining weight, accompanied by a disturbed perception of one’s own physical state (1). Whilst the current literature agrees on an extensive biological foundation underlying the disorder, its nature has not yet been fully understood (2).

Resting-state functional magnetic resonance imaging (rsfMRI) (3) provides important insights into organization and alteration of functional brain networks. In contrast to task-based fMRI, rsfMRI is relatively simple to perform, easily scalable and able to capture information on the brain-network level independent of individual compliance and performance, thus promising to extend the current knowledge on neural alterations in AN (4). Previous rsfMRI research in AN led to a variety of findings, encompassing alterations of inter-as well as intraregional functional connectivity (FC) and activity in executive control, default mode, visual and sensorimotor networks as well as prefrontal and insular cortices (5–10). These alterations may underlie AN-related phenomena such as excessive cognitive control, disintegration of sensory and interoceptive information and persistent rumination about topics related to bodyweight and -shape (5).

The often severe starvation which patients experience in the acute state of AN can cause various somatic and psychiatric symptoms, that are not necessarily a specific product of the disorder but of starvation in general (2). The physical adaptation to starvation may be partly regulated by leptin, a hormonal marker of adiposity and energy storage (11). As was shown for grey matter volume (GMV) during different weight-recovery states of AN (12,13), causes and consequences of undernutrition can influence resting-state brain functioning (14,15). Longitudinal studies are crucial to better understand the development of rsfMRI alterations during the clinical course of AN and to disentangle the contribution of starvation and weight restoration from potential specific AN-traits. Only two studies to date adapted such a longitudinal design providing controversial results on FC alterations (16,17). Whilst Cha et al. (16) reported FC differences to disappear after short-term weight restoration, Uniacke et al. (17) found some evidence for persistent FC alterations. In line with the latter, alterations in executive control (18), default mode (19) and visual networks (20,21) were reported in long-term recovered patients. However, these findings were not consistently replicated, (5,6) may have been confounded by differences in underlying grey matter (22) and were questioned by a recent larger-scale study (23) reporting a normalization of most alterations found in acute AN (7).

To date, due to widespread and inconsistent findings obtained by a variety of data analytic methods with mostly cross-sectional approaches, the question on the nature (e.g., relation between intra- and interregional FC alterations), confounds (e.g., influence of GMV changes) and state- or trait-dependency (influence of acute malnutrition) of rsfMRI alterations in AN remains open.

To address these questions, we examine rsfMRI data in patients in the underweight, short-term weight-restored and long-term recovered state of AN. Given the widespread nature of previous findings, we follow a data-driven approach testing for alterations of whole-brain functional connectivity and activity by utilizing Network Based Statistics (24), Global Correlation (25), Integrated Local Correlation (26) and the fractional Amplitude of Low Frequency Fluctuations (27). We further evaluate if rsfMRI alterations persist when controlling for underlying GMV. Using this integrative approach, we aim to provide valuable insights into the development of resting-state alterations over the clinical course of AN.

## Methods

### Participants

In total, 87 women underwent rsfMRI scanning in two separate cohorts (Table 1; tables S1-3). The first cohort (“acu”) comprised 22 adolescent inpatients with AN (*N* = 1 binge eating/purging subtype) who were scanned at admission (T1acu) and after discharge from treatment (T2acu, *N* = 21) aside with 22 age-matched healthy controls (HCacu; scanned once). The second cohort (“rec”) consisted of 21 young adult patients recovered from adolescent-onset AN for at least 12 months (T3rec) and 22 age-matched HC (HCrec). Except for three patients with AN, the two cohorts comprised separate participants. To avoid any additional assumptions on the data, the cohorts were assumed independent. AN was diagnosed according to DSM-5 criteria (1) and all patients received inpatient treatment at the eating disorders (ED) unit of the Department of Child and Adolescent Psychiatry and Psychotherapy, University Hospital Aachen, Germany. For additional details on the study sample as well as recruitment and scanning procedure, see supplementary methods.

**Table 1:**
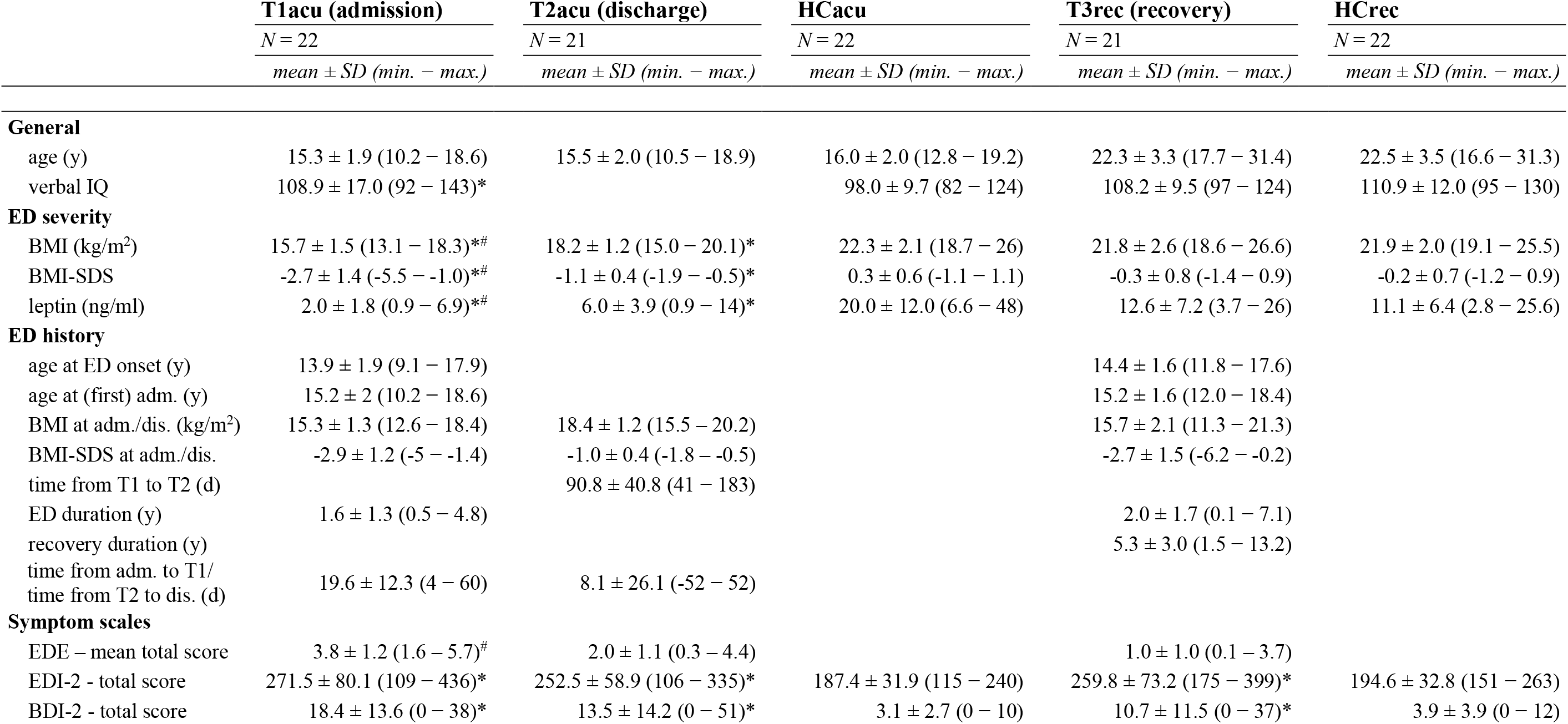
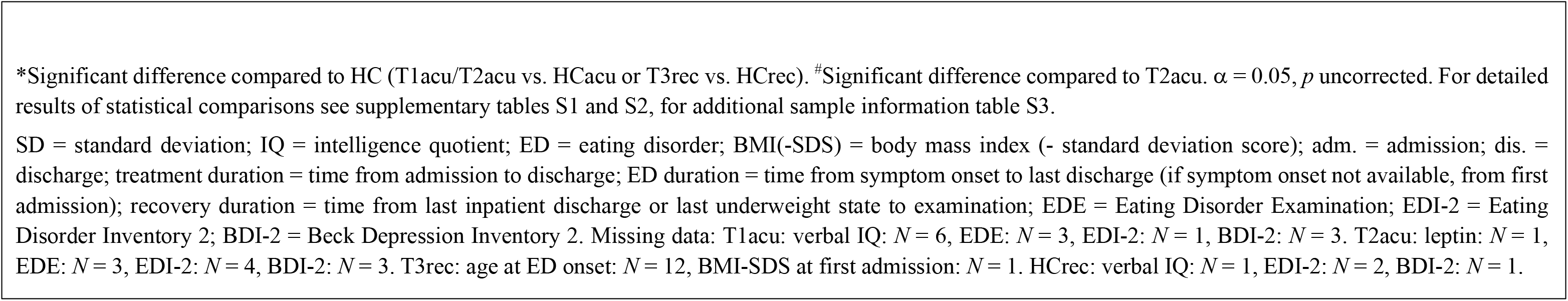
Demographic and clinical sample characteristics.

Both studies were approved by the local ethics committee (cohort 1: EK081/10, cohort 2: EK213/15) and were conducted in accordance with the Declaration of Helsinki. All participants and their legal guardians (if underage) gave written informed consent.

### Clinical Assessments

Current eating disorder diagnosis and severity were examined using the Eating Disorder Examination (EDE) (28) and the Eating Disorder Inventory Revised (EDI-2; 6-point-scale, 11 subscales) (29). Undernutrition was quantified by age- and sex-adjusted BMI standard deviation scores (BMI-SDS) (30,31) and plasma leptin. The sex-adjustment of BMI scores was relevant to evaluating the participant’s nominal underweight (see supplementary methods). Depressive symptoms were assessed with the Beck Depression Inventory (32). Screening for psychiatric comorbidities was conducted using the Kiddie Schedule for Affective Disorders and Schizophrenia (33) or the Mini International Neuropsychiatric Interview (34). The Multiple-choice Vocabulary Test (35) was utilized to approximate verbal intelligence quotient (IQ). Further information on psychopathology and past inpatient treatments were obtained from clinical records. Interviews were conducted by specifically trained and supervised clinical researchers with background in medicine or psychology.

### MRI Data Acquisition

All participants were scanned at the same 3T MRI scanner (Magnetom Prisma; Siemens, Erlangen, Germany). Structural T1-weighted images were acquired using a Rapid Acquisition Gradient Echo (MP-RAGE) sequence with following parameters: cohort 1: TR = 1880 ms, TE = 3.03 ms, Field of View (FoV) = 256 × 256 mm; cohort 2: TR = 2300 ms, TE = 2.98 ms, FoV = 256 × 240 mm; both cohorts: number of slices = 176, voxel size = 1 × 1 x 1 mm^3^. For rsfMRI, all probands underwent a 7 minutes eyes closed T2*-weighted gradient-echo Echo Planar Imaging (EPI) protocol with TR = 2000 ms, TE = 28 ms, flip-angle = 77°, FoV = 192 × 192 mm, number of slices = 34, number of volumes = 210, voxel-size = 3 × 3 x 3.5 mm^3^. All subjects in cohort 1 (patients and HC) were scanned during daytime while continuing their usual food intake or refeeding plans, all subjects in cohort 2 were scanned in the morning after an overnight fasting.

### Preprocessing

Preprocessing of functional and structural images was conducted with SPM12 (36) in a MATLAB (37) environment. For further processing and analyses of rsfMRI data CONN18b (25) was used.

Preprocessing included removal of the first four frames, realignment for motion correction and co-registration to structural images with subsequent spatial normalization into Montreal Neurological Institute space using parameters derived from structural data and interpolation of the data to a 3 mm isotropic resolution. The normalization parameters were also applied (with modulation) to segmented grey matter probability maps to obtain corresponding voxel-wise GMV. A grey matter mask (probability of grey matter > 0.2) was applied to all images to restrict analyses to grey matter tissue. A Gaussian smoothing kernel of 6 mm full-width at half maximum (FWHM) was applied to rsfMRI data. Twenty-four motion parameters (38) aside with mean white matter and cerebrospinal fluid signals were regressed out of the functional data. The resulting images were linearly detrended and temporally band-pass-filtered (0.01 to 0.08 Hz). One HCacu had to be excluded from analysis because of data processing errors. Groups did not differ in regard to mean frame wise displacement (FWD) and no subject exceeded a maximum FWD (translation) of 3 mm (table S4).

### Analyses of demographic and clinical data

Statistical analyses of demographic and clinical data were conducted using R (39) and *jamovi* (40). *T*-, Mann-Whitney-*U*- or Wilcoxon-tests were used as appropriate to compare characteristics between groups.

### Primary analyses of rsfMRI data

Primary rsfMRI analyses consisted of (I) calculation of resting-state FC and activity measures based on whole-brain voxel-wise data and (II) a parcellation-based approach applying Network-Based Statistics (NBS) (24). The derived measures were compared between AN- and corresponding HC-groups (T1acu vs. HCacu, T2acu vs. HCacu, T3rec vs. HCrec) and between T1acu and T2acu to assess temporal evolution of rsfMRI alterations in the acute-recovery phase. Additionally, an “interaction” design including acute and recovered patients and corresponding control groups was applied testing for group-by-time interactions between patients at admission and in long-term recovery. Extraction of voxel-wise rsfMRI measures, calculation and visualization of NBS results were conducted using CONN.

Global Correlation (GC) was calculated as the average of bivariate correlations between the Blood oxygenation level dependent (BOLD) signal of a given voxel and every other voxel (25). Integrated Local Correlation (LC) was computed as the average bivariate correlation between each voxel and its neighboring voxels weighted by a Gaussian convolution with 6 mm FWHM (26). Fractional Amplitude of Low Frequency Fluctuations (fALFF) was calculated at each voxel as the root mean square of the BOLD signal amplitude in the analysis frequency band (0.01 – 0.08 Hz) divided by the amplitude in the entire frequency band (27). Group comparisons were conducted using t-contrasts within general linear models while controlling for age. Data from T1acu were compared to T2acu using a repeated measures design. An exact permutation based (1000 permutations) cluster threshold (*p* < 0.05) was applied in all analyses combined with an uncorrected voxel-wise threshold of *p* < 0.01 (22) allowing for an accurate control of the false positive rate while maintaining sensitivity to potentially weak resting-state alterations (41).

For NBS analyses, preprocessed functional images were (without smoothing) parcellated into 100 cortical (42) and 16 subcortical brain regions (Neuromorphometrics, Inc.). For reporting, Automated Anatomical Labeling atlas (43) regions corresponding to the centroid coordinates of the “Schaefer-atlas” were used. BOLD signal time courses were averaged within each region of interest (ROI). Subject-wise bivariate correlation matrices (116 × 116) were calculated and Fisher’s *z*-transformed. Each ROI-to-ROI connection was compared between groups and resulting *p*-values were thresholded at an uncorrected level of *p* < 0.01. Within this set of supra-threshold connections, all possible connected (sub-)networks were identified. Using permutation testing (10000 iterations), resulting subnetworks were assessed for statistical significance based on their sizes while controlling the family-wise error rate (24).

In additional sensitivity analyses, contrasts yielding significant group differences were recomputed by including mean rotational and translational FWD as additional covariates to control for motion at the group level.

### Post-hoc group comparisons

Implementation and visualization of post-hoc comparisons and correlation analyses were conducted using *jamovi* an*d* R. To better understand group differences, FC of every connection included in significant NBS subnetworks was averaged for each subnetwork (separately for increased and decreased connections) and values of every voxel included in significant clusters were averaged per cluster. These measures representing whole networks or clusters, respectively, were used for further analyses. Age was controlled for in all post-hoc analyses. The correction for multiple comparisons inherent to the NBS procedure does not allow for inference about individual connections (24). To explore the neuroanatomy of the NBS results, the region with the strongest alteration in node degree (the region showing the largest number of altered connections) in each subnetwork was identified. Connections between these regions and any other ROI that showed significant differences (Bonferroni-corrected) between groups were identified by applying a seed-to-ROI approach.

To illustrate the temporal development of resting-state properties that were altered in acute AN, averaged values from the T1acu < HCacu and T1acu < T2acu contrasts were compared between AN and corresponding HC using analyses of covariance (ANCOVAs) or paired *t*-tests for each network or cluster. To demonstrate that altered functional connections and clusters coinciding between different contrasts showed consistent recovery dynamics, overlapping FC and voxel-wise values were extracted and compared as stated above. Bonferroni correction was applied to control for the number of tests per rsfMRI modality and contrast. To test for statistical biases and to allow for an appropriate interpretation of the results when comparing data based on the T1acu vs. HCacu comparison between T2acu and the same control group, we performed a simulation experiment. Results indicated that the likelihood of observing significant between-group differences increases for such comparisons (see supplementary methods, results and figure S3). In additional sensitivity analyses, we explored the effects of controlling for IQ, for time between inpatient admission and T1 (to control for delayed scanning) and for BMI-SDS as well as of excluding participants taking psychoactive medication. To evaluate the potential impact of outliers on rsfMRI group differences, we recalculated corresponding post-hoc group comparisons using non-parametric statistics (see supplementary methods and results).

### Relationships among rsfMRI measures

We further assessed how the different rsfMRI alterations identified in acute AN relate to each other. For this, Pearson correlations between all rsfMRI measures in AN at T1 as well as between changes in rsfMRI measures from T1 to T2 were computed (false discovery rate corrected).

### Relationship to clinical outcomes

Next, we tested for associations between rsfMRI results and clinical variables. Pearson correlations (Bonferroni corrected) were computed between rsfMRI alterations (T1acu vs. HCacu and delta T2-T1) and BMI-SDS, leptin, total scores on EDE, EDI-2 and BDI-2, the time between eating disorder onset and inpatient admission (T1acu) as well as the time from T1 to T2 (delta T2-T1).

### Effects of grey matter volume on rsfMRI alterations

To evaluate the impact of GMV on rsfMRI alterations, voxel-wise GMV was averaged for each cluster- or subnetwork from resting-state analyses and compared between groups using ANCOVAs or paired *t*-tests where appropriate. To evaluate influences of GMV changes on NBS results, group comparisons were recomputed after GMV was regressed out of the averaged network FC (separately for acute and recovered cohorts). For voxel-wise rsfMRI measures, individual GMV was regressed out at each voxel included in significant clusters. Cluster values were then averaged and compared between groups. To assess the impact of GMV correction on the magnitude of resting-state group differences, Cohen’s *d* values (with Hedge’s *g* correction) were computed for all significant rsfMRI measures before and after controlling for GMV (aside with respective GMV effect sizes).

## Results

### Demographic and clinical sample characteristics

AN and corresponding HC did not differ regarding age. In cohort 1, AN showed higher verbal IQ compared to HC. Patients from the two study cohorts were comparable in regard to age at ED onset as well as age and BMI-SDS at first inpatient admission. As expected, patients with AN at T1 showed significantly lower BMI (-SDS) and leptin levels and higher scores on EDE, EDI-2 and BDI-2. Average BMI increase during inpatient treatment was 20.8 ± 9.5%. T3rec patients exhibited elevated EDI-2 scores (Table 1; tables S1 and S2).

### Parcellation-based results

Using NBS, we identified significant differences between T1acu and HCacu and between T1acu and T2acu (Figures 1A and C; figure S1, table S5). Included connections were largely decreased for T1acu constituting a widespread network across the whole brain. T2acu as well as T3rec patients did not differ from HC. Post-hoc analyses based on T1acu findings revealed smaller but significant FC reduction for T2acu but not for T3rec (Figures 1E-G; table S6).

**Figure 1:**
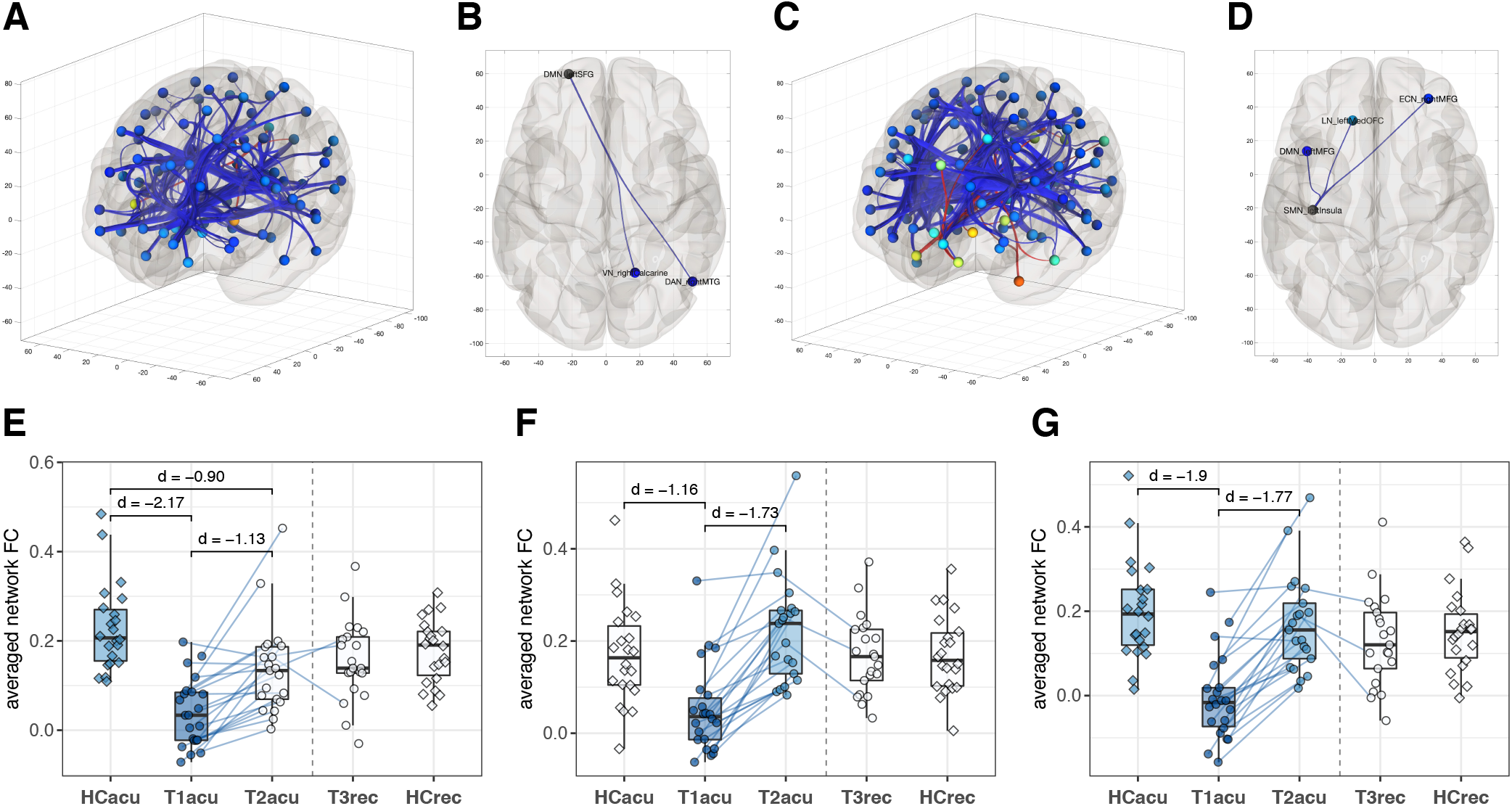
Network Based Statistics. A: NBS subnetwork resulting from T1acu vs. HCacu comparison. B: Seed-to-ROI: Within the T1acu vs. HCacu subnetwork, functional connectivity (FC) between left superior frontal gyrus (seed) and right calcarine sulcus/ middle temporal gyrus shows the strongest reduction (*p* < 0.05/115). C: NBS subnetwork resulting from T1acu vs. T2acu comparison. D: Seed-to-ROI: Within the T1acu vs. T2acu subnetwork, FC between left insula (seed) and prefrontal regions (middle frontal and orbitofrontal gyri) shows the strongest reduction. E: Post-hoc comparison of averaged T1acu < HCacu subnetwork FC. F: Post-hoc comparison of averaged T1acu < T2acu subnetwork FC. G: Post-hoc comparison of subnetwork FC averaged from connections overlapping between T1acu < HCacu and T1acu < T2acu subnetworks. Boxplot figures represent post-hoc comparisons of results from T1acu < HCacu and T1acu < T2acu contrasts. Each box represents one group, the scatter points represent single subjects (AN-subjects as circles, HC-subjects as squares). Blue lines indicate matching values of individual AN-subjects. The groups involved in the original, “primary”, comparisons are colored to highlight circular statistical tests. Only groups within each study cohort were compared to each other, the dashed line separates the cohorts. Significant group comparisons (Bonferroni-corrected) are marked with brackets and complemented by corresponding effect sizes (Cohen’s *d*, Hedge’s *g* corrected).

Left anterior superior frontal gyrus (SFG; T1acu vs. HCacu subnetwork; figure S2A) and left insular cortex (T1acu vs. T2acu subnetwork; figure S2B) displayed the largest decreases in node degree. Using these seeds, we found decreased FC for T1acu between left SFG and ROIs in the left medial temporal gyrus (MTG) and right calcarine sulcus as well as between left insula and bilateral medial frontal and left medial orbitofrontal cortex (Figures 1B and D; table S5). NBS results were largely similar when additionally controlling for FWD, IQ, admission-scan-delay and BMI-SDS, when excluding participants taking psycho-active medication or when using non-parametric statistics (tables S7-S12).

### Voxel-wise results

In pairwise comparisons, significant differences were observed for all voxel-wise resting-state measures for T1acu < HCacu and T1acu < T2acu contrasts (Figures 2A-C, brain slices; table S13). No other contrasts were significant. T1acu patients exhibited reduced GC in bilateral prefrontal regions in comparison to HC and in the right insula relative to T2acu (Figure 2A). In both contrasts, GC was reduced in bilateral sensorimotor areas. LC was reduced in T1acu in bilateral sensorimotor areas, right SFG and bilateral precuneus relative to HCacu and in bilateral sensorimotor, right fusiform and left MTG areas relative to T2acu (Figure 2B). We found a significant interaction in the medial posterior frontal cortex for LC comparing T1acu and T3rec patients relative to respective HC (Figure 2B). Fractional ALFF was reduced for T1acu in bilateral precuneus and calcarine sulcus compared to T2acu and HCacu, in left parietal regions relative to HCacu and in left MTG relative to T2acu (Figure 2C).

**Figure 2:**
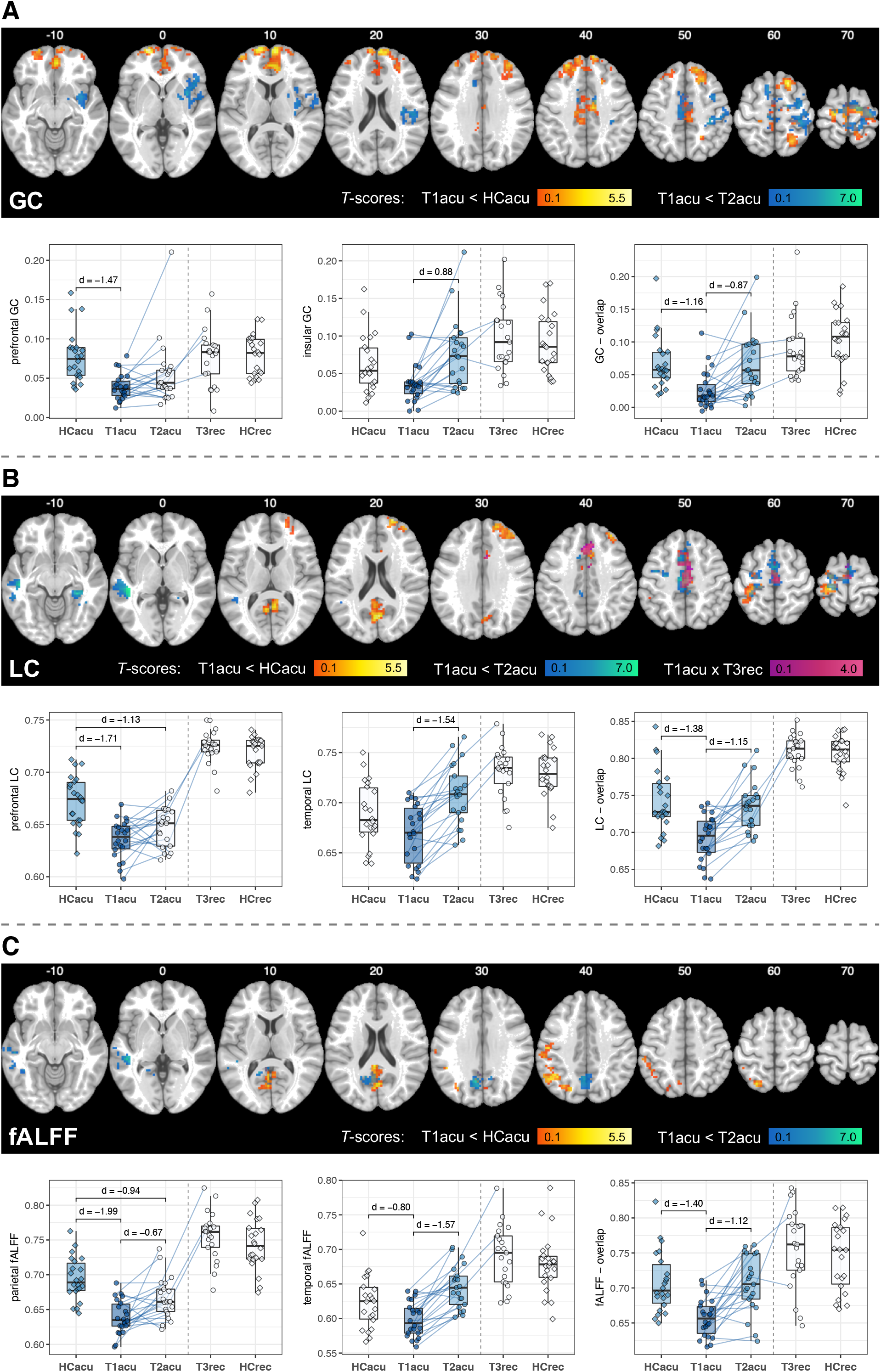
Voxel-wise functional connectivity and activity. Brain slices: Clusters of significantly reduced voxel-wise resting-state measures in T1acu patients compared to HCacu (red-yellow) and compared to T2acu (blue-green); results from T1acu < HCacu x T3rec > HCrec contrast are displayed in purple. Color brightness reflects T-value size at a given voxel. Boxplot diagrams: Left and middle: Clusters with the largest effect sizes in post-hoc comparisons of each voxel-wise resting-state measure. Right: Post-hoc comparisons of clusters overlapping between T1acu < HCacu and T1acu < T2acu contrasts. For general descriptions of the boxplot diagrams, refer to the legend of Figure 1. A: Global Correlation (GC). Alterations of prefrontal (T1acu < HCacu) and right insula GC (T1acu < T2acu) show the larges effect sizes. B: Integrated Local Correlation (LC). The larges effect sizes were observed for LC in right superior frontal gyrus (T1acu < HCacu) and in left middle temporal gyrus (T1acu < T2acu). C: Fractional Amplitude of Low Frequency Fluctuations (fALFF). Left parietal fALFF (T1acu < HCacu) and fALFF in left middle temporal gyrus (T1acu < T2acu) display the larges effect sizes.

From all identified cluster per modality, (right) prefrontal GC and LC and left parietal fALFF showed the largest effect sizes (Figures 2A-C, left boxplot diagrams; Figure 3C; table S6). In post-hoc group comparisons of these clusters, right prefrontal LC and left parietal fALFF were also reduced in T2acu compared to HCacu (Figures 2B and C, left boxplot diagrams; table S6). Brain areas coinciding between the significant contrasts showed reductions for T1acu compared to both, HCacu and T2acu, for all modalities (Figures 2 A-C, right boxplot diagrams; table S6). All results remained largely similar when controlling for FWD, IQ and admission-scan-delay or when excluding medicated participants (tables S14, S8, S9, S11). When controlling for BMI-SDS in the T1acu < HCacu contrast, only alterations in prefrontal GC and LC and left parietal fALFF remained significant; in the T1acu < T2acu contrast, all differences, except for sensorimotor GC, remained significant (table S10). Outliers in the data did not have significant impact on group effects (table S12).

**Figure 3:**
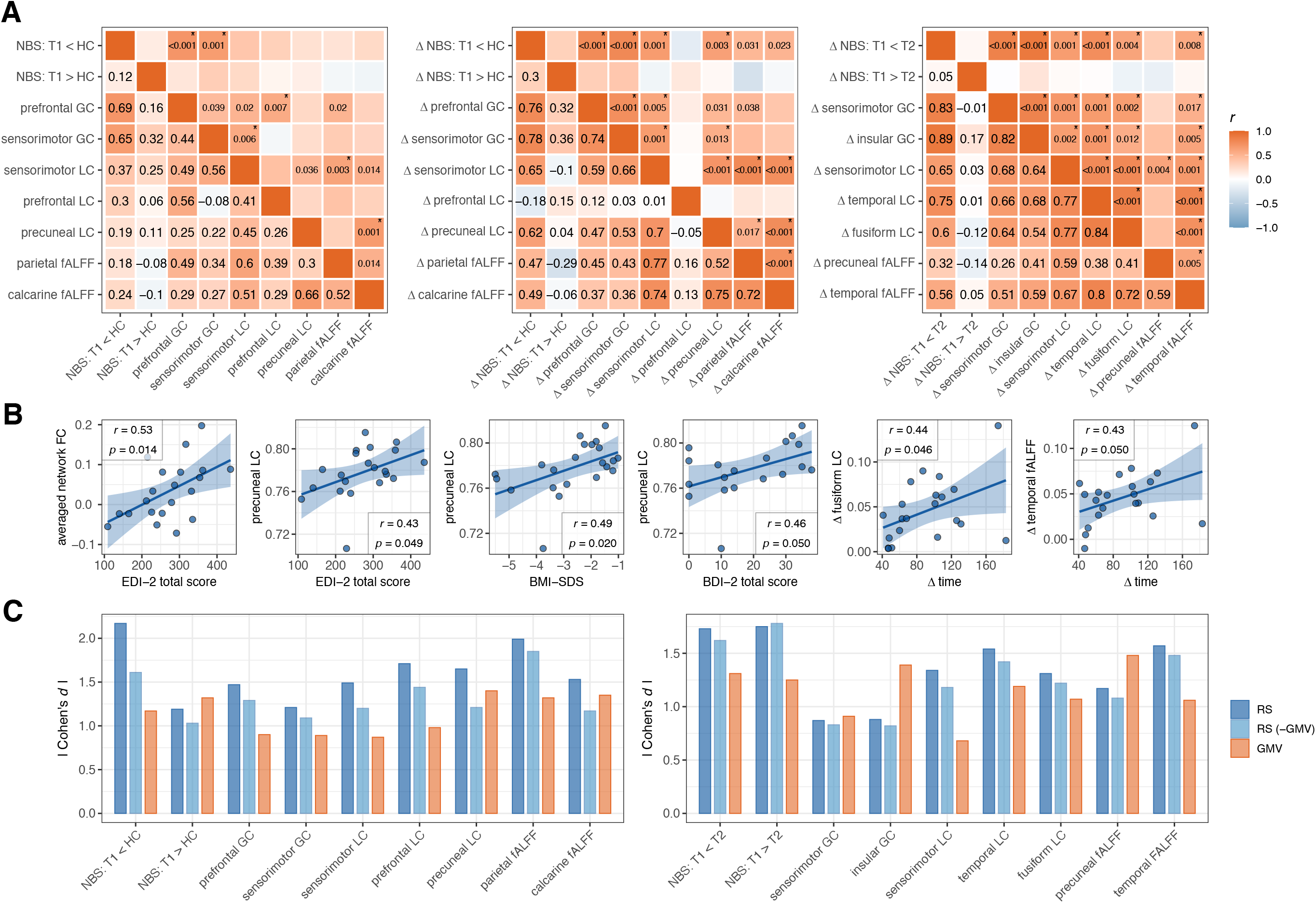
Correlations among resting-state measures and with clinical outcome; Effect sizes of resting-state group comparisons with and without controlling for voxel-wise grey matter volume. A: Left: Heatmap representing correlations of rsfMRI results from T1acu vs. HCacu comparison across resting-state measures in the T1acu group. Middle: Correlations between T1-T2-changes (“delta”) in rsfMRI results from T1acu vs. HCacu comparisons. Right: Correlations between T1-T2-changes in rsfMRI results from T1acu vs. T2acu comparisons. Colors of single squares represent the Pearson correlation coefficient *r*. The left lower triangle shows Pearson’s *r* values, the right upper triangle shows associated *p*-values (if *p* > 0.05). * Correlations that remained significant after correcting for the false discovery rate (Benjamini-Hochberg procedure). B: 1 - 4: Associations between results from T1acu vs. HCacu rsfMRI comparisons and clinical variables in the acute AN group. 5 & 6: Associations between changes in resting-state measures and changes in clinical variables from T1 to T2. Since no correlation survived Bonferroni-correction, correlations have to be considered exploratory. Pearson’s *r* and uncorrected *p*-values are shown. Scatter points represent patients with acute AN at T1, respectively differences between T1 and T2 (“delta”). Blue lines display the fitted linear regression function, blue areas the corresponding 95% confidence interval. C: Effect sizes (Cohen’s *d*, Hedge’s *g* corrected) from post-hoc comparisons of resting-state measures (RS), resting-state measures after voxel-wise regression of grey matter volume (RS-GMV) and grey matter volume (GMV). Left: T1acu vs. HCacu; right: T1acu vs. T2acu comparisons. NBS = Network Based Statistics; GC = Global Correlation; LC = Integrated Local Correlation; fALFF = fractional Amplitude of Low Frequency Fluctuations; FC = functional connectivity; EDI-2 = Eating Disorder Inventory 2; BMI-SDS = body morph index – standard deviation score; EDE = Eating Disorder Examination; T1 = T1acu; T2 = T2acu; HC = HCacu.

### Correlations among rsfMRI measures

Considering the multitude of identified alterations, we explored which of the observed rsfMRI changes are related to each other using correlation analyses. We found strong positive correlations between measures capturing global (NBS, GC) respectively local (LC, fALFF) resting-state characteristics and between topologically close alterations obtained by different methods (e.g. GC and LC in prefrontal or motor areas). Changes in rsfMRI measures from T1 to T2 correlated strongly across all measures, except for T1acu > HCacu and T1acu > T2acu network FC and prefrontal LC (Figure 3A).

### Relationship to clinical outcomes

No correlations between rsfMRI results and clinical variables remained significant after correction for multiple comparisons (table S15). At an uncorrected p < 0.05, in patients with acute AN T1acu < HCacu network FC was positively associated with EDI-2 scores. Bilateral precuneal LC was positively correlated with BMI-SDS, EDI-2 and BDI-2 in patients with acute AN. Changes in right fusiform LC and left temporal fALFF correlated positively with time from T1 to T2 (Figure 3B). Given the unexpected direction of correlations with global ED severity scores, we performed additional exploratory correlation analyses that suggested strongest correlations between FC changes and EDI-2 subscales such as maturity fear and social insecurity rather than core eating behavior abnormality subscales (supplementary results, figure S4).

### Effects of grey matter volume on rsfMRI alterations

GMV was significantly reduced in acute AN in all rsfMRI clusters and networks (Figure 3C; table S16). When regressing GMV out of voxel-wise GC, LC and fALFF maps or averaged network FC, group differences and corresponding effects sizes remained largely unchanged for all measures (Figure 3C; table S17). GMV did not correlate with any of the rsfMRI measures (table S18).

## Discussion

We systematically explore whole-brain rsfMRI alterations in acute, short-term and long-term recovered, adolescent-onset AN. In acute patients, we find spatially widespread decreases of local and global FC and local activity measures. These alterations are independent of underlying GMV and normalize largely with short-term weight restoration to being absent in the long-term recovered group.

In line with several previous publications, we find largely decreased intra- and interregional rsfMRI measures in acute AN in bilateral prefrontal, sensorimotor, left parietal, left temporal, bilateral precuneal and insular regions (7–10,20,21,44–48). In contrast to some previous studies, we do not find increases of intraregional (7) or interregional FC and activity (16,17,49–51). This discrepancy may be due to differences in sample size, methodology as well as in most cases shorter illness duration and younger age of our sample (table S19).

We find a complete normalization of all rsfMRI alterations in long-term recovered patients with a similar trend observed immediately after refeeding therapy. These findings are in line with results of Seidel et al. (23) and Cha et al. (16) and are further supported by similar conclusions from longitudinal task-based fMRI studies (52,53). This lack of differences between long-time recovered AN and HC in our study further supports the notion that the identified rsfMRI alterations may constitute temporal, presumably starvation-related, state effects of the disorder (23). Differences to previous research indicating persistent alterations in varying brain networks in short-(17) and long-term recovered AN (18–21) may be due to the longer average recovery time (5.3 ± 3 years) and the considerably shorter illness durations in our cohorts (tables S20, S21). This further underlines the importance of early identification and treatment of AN during adolescent age (2).

Whereas underweight-related GMV decreases in AN were consistently shown (12,13), the dissociation between functional (resting-state) and structural alterations has not been systematically addressed. Our results are supported by preliminary data suggesting decreased function-structure relation (7) and GMV-independency of resting-state alterations in AN (21). The finding of a global reduction in FC, the normalization of rsfMRI alterations with recovery and the absence of robust clinical correlations support the notion of rsfMRI alterations being starvation-related phenomena. The missing relationship between resting-state recovery and increasing BMI as well as GMV changes during inpatient treatment may possibly be explained by a mechanism of recovery proceeding faster than weight-restoration such as, for example, short-term changes in energy-supply (14,15). The near absence of resting-state alterations combined with persisting reduced bodyweight and elevated ED psychopathology at inpatient discharge supports this hypothesis. However, considering the moderate sample size in our study, AN psychopathology may still have an independent influence on recovery of resting-state brain functioning.

Alterations of bilateral prefrontal and left parietal rsfMRI measures display the largest effect sizes in acute AN in our study. Both regions play a central role in altered cognitive control associated with AN-pathophysiology as indicated by brain imaging, theoretical and behavioral research (5,6,54,55). Alterations in parietal regions in AN have repeatedly been linked to distorted body image (56). Simultaneously reduced local and global connectivity in prefrontal brain regions point to a common biological background (57,58). In line with that, we detect significant associations between topologically close alterations in different measures and between measures capturing local, respectively global rsfMRI characteristics. In contrast, correlational analyses between changes from T1 to T2 show a much denser pattern of significant correlations across all measures. These findings indicate a global trend of recovery with no notable emphasis on certain resting-state alterations or specific metrics.

As stated above, we do not find correlations between rsfMRI and clinical measures surviving correction for multiple comparisons. Considering the small sample size and the adopted exploratory approach this is not surprising. Nonetheless, we find some preliminary evidence for associations between rsfMRI and clinical measures including an unexpected positive correlation between ED severity scales and FC in acute AN that would require replication in larger samples.

### Limitations

A major limitation of the study is the relatively small sample size of the cohort evaluated here preventing detection of potentially weaker rsfMRI alterations and limiting the generalizability of our findings (59). Furthermore, the AN-, respectively starvation-related differences of pubertal status between acute patients and controls and the broad age range of the acute patient group could influence rsfMRI group differences (60,61). Despite these limitations, considering the lack of previous longitudinal rsfMRI studies in AN, our results provide a basis for future hypothesis-driven and replication studies to build upon.

## Conclusion

We provide novel insight into extent and recovery of resting-state brain functioning during the clinical course of AN. Resting-state alterations in AN are independent of GMV and are compatible with starvation-related phenomena indicating their potential as state-markers of the disorder. Consistent with clinical findings, brain regions previously associated with cognitive control and body image disturbance show the most pronounced alterations in acute AN. Absence of these alterations in our fully recovered group with relatively short illness duration underlines the importance of early identification and treatment in AN.

## Data Availability

The individual data are available on request. Statistical group-maps were uploaded to NeuroVault.

## Acknowledgements

The study was supported by the Swiss Anorexia Nervosa foundation (cohort 2; Grant number: 53-15). A preprint of this manuscript is available from medRxiv (https://doi.org/10.1101/2020.06.21.20135566). Statistical group-level maps derived from the reported analyses were uploaded to NeuroVault (https://neurovault.org/collections/EIFQNRMC/).

## Author’s contribution

LDL performed all analyses and wrote the manuscript with support of JD and GvP. LDL, KK, SBE, JS and JD designed the overall study. JO, KB, LS and LDL conducted the clinical studies and gathered the data under supervision of JS and KK. All authors reviewed and commented on the manuscript.

