## Supplementary material for "Recovery-associated resting-state activity and connectivity alterations in Anorexia nervosa"

### Supplementary Methods

#### Recruitment procedure and additional sample descriptions

Only patients meeting the following criteria were included in the acute Anorexia nervosa (AN; T1/T2acu) group: (I) Diagnosis of AN according to DSM-V criteria (1), (II) inpatient admission to specialized eating disorder (ED) treatment at the Department of Child and Adolescent Psychiatry and Psychotherapy of the University Hospital Aachen, Germany, (III) body mass index (BMI) below the $10^{th}$ age- and sex-adjusted percentile. Patients were included in the recovered patients (T3rec) group if (I) they had received a past AN diagnosis and treatment as stated above, (II) had a current BMI over 18.5 kg/$m^2$ or the $10^{th}$ percentile (if underage), (III) did not fulfill criteria of any ED diagnosis according to a standardized clinical interview (2) at the time of testing, (IV) nor were underweight in the last 12 months or (V) exhibited current amenorrhea. Only subjects with (I) a BMI over 18.5 kg/$m^2$ or the $10^{th}$ percentile and (II) without any history of psychiatric or neurological disorders or psycho-active medication use were included in the control groups (HCacu/ HCrec). Additionally, all subjects had to exceed an intelligence quotient (IQ) of 80 and fulfill functional magnetic resonance imaging safety criteria. Anorexia nervosa patients were recruited from the Department of Child and Adolescent Psychiatry and Psychotherapy, University Hospital Aachen, Germany. Healthy controls were reached via participation in former studies, flyers and social media from the surrounding of the RWTH Aachen.

In total, we collected resting-state functional magnetic resonance imaging (rsfMRI) scans of 102 subjects. Applying the listed criteria, we excluded 15 subjects from the final analyses: $N$ = 4 HCacu (2 over-threshold ED-scores, 3 BMI criteria, 1 data processing errors), $N$ = 8 T3rec (5 acute ED diagnosis, 2 BMI criteria, 1 acute suicidal tendencies), $N$ = 3 HCrec (2 BMI criteria, 1 anti-depressive medication).

Since not all patients (*N* = 5 below the $10^{th}$ sex- and age-adjusted percentile) reached normal-range BMI levels at discharge, the complete group was considered only partially, “short-term”, weight-recovered.

All patients in cohort 1 (patients and controls) were scanned at varying times over the day (ranging from 9 to 10 am to afternoon) while continuing their usual food intake or refeeding treatment plans. Calorie intake per day ranged from 1200 to 2600 kcal in the T1acu group and from 2400 to 3000 kcal in the T2acu group, allocated to 6 meals per day and depending on the food intake prior to inpatient admission. All participants in cohort 2 (patients and controls) were scanned in the morning (usually beginning at 8 to 10 am) after an overnight fasting.

**Assaying of plasma leptin**

To quantify plasma leptin levels, blood samples were drawn preceding the fMRI scan, centrifuged, aliquoted, stored at -80°C and analyzed with an Enzyme Linked Immunosorbent Assay kit (ELISA; Mediagnost, Reutlingen, Germany).

**Used software, toolboxes and packages**

In addition to those cited in the main text (3–7), following software and R-packages were used to create visualizations: *MRIcron* (8), *ggplot2* (9), *ggsignif* (10), *ggcorrplot* (11), *patchwork* (12) and *cowplot* (13).

**Simulation experiment to assess potential statistical bias in post-hoc comparisons**

In post-hoc analyses, we systematically explored the temporal development of rsfMRI alterations identified in T1acu < HCacu and T2acu contrasts by comparing the results between T2acu (and T3rec) and HCacu. The latter may be affected by statistical bias resulting from comparing data based on T1acu-HCacu-differences between T2acu and the same controls. To assess the effect of this potential bias, we conducted a simulation experiment. Three vectors of random normally distributed numbers (x, y, z; resembling data from T1acu, T2acu and HCacu) were generated, while accounting for the correlation between x and y. When a *t*-test comparing x to z resulted in a significant result ($p < 0.05$), a second *t*-test comparing y to z was conducted. The process was repeated 100.000 times and the distribution of *p* values was plotted in figure S3. The MATLAB code to generate the *p* distribution is provided below.

```
p_all = [];
rng(1);
for i = 1:100000
    xy = mvnrnd([0 0], [.9 .4; .4 .9], 21);
    z = randn(22, 1);
    [h, p1] = ttest2(xy(:,1), z);
    if p1 < .05
        [h, p2] = ttest2(xy(:,2), z);
        p_all(end+1,:) = [p1 p2];
        clear('p2');
    end
end
```

**Sensitivity analyses**

To control for motion artifacts at the group level, we averaged frame-wise displacement (FWD) parameters to two scores representing mean translation and rotation FWD. These were included as additional covariates in group-level analyses of rsfMRI data.

We furthermore recomputed all post-hoc-comparisons that showed significant group differences by (I) including verbal IQ as covariate (T1/T2acu vs. HCacu) to assess whether differing IQ values observed in cohort 1 influenced rsfMRI group differences; (II) including the time between inpatient admission and T1acu as covariate (T1acu vs. T2acu) to control for the impact of delayed scanning, (III) including BMI-standard deviation score (BMI-SDS) as covariate in all comparisons to assess the extent to which resting-state alterations in AN were mediated by starvation and (IV) excluding participants taking psychoactive medication at the time of scanning from all comparisons. For this, (repeated measures) analyses of covariances (RM-ANCOVAs) as well as paired *t*-tests were used.

Finally, we assessed all averaged resting-state measures for outliers (defined as values exceeding 2 standard deviations in relation to the group-wise mean) and recomputed corresponding significant post-hoc group comparisons using Wilcoxon or Mann-Whitney-*U*-tests.

**Additional correlation analyses**

Counterintuitively, we observed a strong positive association between T1acu < HCacu network functional connectivity (FC) and self-reported eating disorder symptom severity (Eating Disorder Inventory 2 (EDI-2) total score). Precuneal Integrated Local Correlation (LC) was also positively related to EDI-2 and furthermore to the Beck Depression Inventory (BDI-2) total score. The EDI-2 is composed of 11 subscales that measure eating disorder specific (e.g. *Drive for thinness*, *Body dissatisfaction*, *Bulimia*) as well as unspecific, partly state-related characteristics (e.g. *Interpersonal distrust*, *Impulse regulation*, *Interoceptive awareness*, *Maturity fears*, *Perfectionism*, *social insecurity*) (14,15). To evaluate whether specific patterns of associations between FC and the various traits and states measured by the EDI-2 emerged, we calculated Pearson correlations between T1acu < HCacu network FC and all EDI-2 subscales. Furthermore, we evaluated how the association developed over time (Pearson correlations in T2acu and T3rec groups) and whether it differed between patients with AN in different states of recovery (ANCOVA with EDI-2 total score as dependent variable; group x network FC interaction). Lastly, we explored if the EDI-2 was related to starvation severity and depressive symptoms in patients with acute AN by calculating Pearson correlations between EDI-2 total score and BMI-SDS, plasma leptin as well as BDI-2 total score.

### Supplementary Results

#### Results of post-hoc group comparison simulation experiment

The above-described simulation experiment resulted in a strongly right skewed *p* value distribution indicating a higher probability of significant effects between y and z (resembling T2acu and HCacu) if a significant effect between x and z (resembling T1acu and HCacu) was observed (figure S3). Transferring these simulated results to our data indicating, in most cases, a normalization of rsfMRI measures at T2 (no significant T2acu < HCacu differences of clusters identified at T1), influence of statistical bias on our findings regarding rsfMRI normalization seems highly unlikely.

#### Impact of potential confounders on rsfMRI results

Including possible confounding variables in group comparisons of rsfMRI results did not introduce major changes to the results. Only BMI-SDS may have a considerable impact on AN-HC-differences in acute patients.

When including FWD in Network Based Statistics (NBS) analyses (T1acu vs. HCacu), the subnetwork size increased and an additional connection between the prefrontal seed ROI and the left calcarine sulcus emerged in the post-hoc seed-to-ROI approach (table S7). When controlling for FWD in the calculation of voxel-wise rsfMRI group differences, original clusters sizes changed slightly, additional clusters of reduced LC (right temporal and orbitofrontal cortices) emerged and the T1acu < HCacu x T3rec > HCrec cluster did not remain significant (table S14).

Whilst controlling for verbal IQ in post-hoc comparisons (T1/T2acu vs. HCacu), only group differences in T1acu > HCacu network FC and bilateral sensorimotor Global Correlation (GC) lost significance (table S8). Note that verbal IQ was available for only $N = 16$ patients with acute AN. Including admission-T1-time as covariate in T1acu vs. T2acu post-hoc comparisons affected significance of GC clusters and left temporal LC (table S9). When including BMI-SDS as covariate in T1/T2acu vs. HCacu comparisons, only differences of bilateral prefrontal GC, right prefrontal LC and left parietal fractional Amplitude of Low Frequency Fluctuations (fALFF) were significant. However, T1acu vs. T2acu differences, except for sensorimotor GC, remained stable (table S10). When excluding participants taking psychoactive medication (T1acu: $N$ = 4, T2acu: $N$ = 6, combined: $N$ = 8), all differences remained significant with in most cases even increasing effect sizes (table S11).

Except for left temporal LC and fALFF in the T1acu vs. T2acu contrast, for all post-hoc comparisons in at least one of the included groups outlier were identified (maximum $N$ = 2). Also using non-parametric statistics, all group differences identified in the main analyses were significant (table S12).

**Exploration of rsfMRI – ED symptom severity correlations**

The only significant (positive) correlations emerged between T1acu < HCacu network FC and the EDI-2 scales *Interoceptive awareness* ($r$ = 0.55, $p$ = 0.010), *Maturity fear*, *Social insecurity* (both $r$ = 0.63, $p$ = 0.002) and *Impulse regulation* ($r$ = 0.45, $p$ = 0.041), but not with eating disorder specific scales (figure S4). In T2acu and T3rec, the correlations with EDI-2 total score were not significant; T2acu: $r$ = 0.20, $p$ = 0.440; T3rec: $r$ = -0.37, $p$ = 0.098. However, the slope of the regression function changed significantly; $F$ = 5.62, $p$ = 0.006. At T1, EDI-2 total score was not related to BMI-SDS ($r$ = 0.03, $p$ = 0.884), nor to plasma leptin ($r$ = -0.07, $p$ = 0.757), but was strongly correlated with BDI-2 total score ($r$ = 0.86, $p$ < 0.001).

### Supplementary Tables

| | T1acu (admission) | T2acu (discharge) | HCacu | Statistics | | | | | |
|---|---|---|---|---|---|---|---|---|---|
| | $N$ = 22 | $N$ = 21 | $N$ = 22 | T1acu vs. HCacu | | T2acu vs. HCacu | | T1acu vs. T2acu | |
| | *mean ± SD (min. – max.)* | *mean ± SD (min. – max.)* | *mean ± SD (min. – max.)* | $T$ or $U$ | $p$ | $T$ or $U$ | $p$ | $T$ or $W$ | $p$ |
| **General** | | | | | | | | | |
| Age (y) | 15.3 ± 1.9 (10.2 – 18.6) | 15.5 ± 2.0 (10.5 – 18.9) | 16.0 ± 2.0 (12.8 – 19.2) | $T$ = -1.23 | 0.227 | $T$ = -0.83 | 0.409 | | |
| verbal IQ | 108.9 ± 17.0 (92 – 143) | *n.a.* | 98.0 ± 9.7 (82 – 124) | $U$ = 106.50 | 0.041* | | | | |
| **ED severity** | | | | | | | | | |
| BMI (kg/m$^2$) | 15.7 ± 1.5 (13.1 – 18.3) | 18.2 ± 1.2 (15.0 – 20.1) | 22.3 ± 2.1 (18.7 – 26) | $T$ = -12.08 | < 0.001* | $T$ = -8.00 | < 0.001* | $T$ = -7.6 | < 0.001* |
| BMI-SDS | -2.7 ± 1.4 (-5.5 – -1) | -1.1 ± 0.4 (-1.9 – -0.5) | 0.3 ± 0.6 (-1.1 – 1.1) | $U$ = 1.00 | < 0.001* | $T$ = -8.95 | < 0.001* | $W$ = 1.00 | < 0.001* |
| leptin (ng/ml) | 2.0 ± 1.8 (0.9 – 6.9) | 6.0 ± 3.9 (0.9 – 14.0) | 20.0 ± 12.0 (6.6 – 48.0) | $U$ = 1.50 | < 0.001* | $U$ = 15.00 | < 0.001* | $W$ = 6.00 | < 0.001* |
| **ED history** | | | | | | | | | |
| age at ED onset (y) | 13.9 ± 1.9 (9.1 – 17.9) | | | | | | | | |
| age at first adm. (y) | 15.2 ± 2 (10.2 – 18.6) | | | | | | | | |
| BMI at adm./dis. (kg/m$^2$) | 15.3 ± 1.3 (12.6 – 18.4) | 18.4 ± 1.2 (15.5 – 20.2) | | $T$ = -13.31 | < 0.001* | $T$ = -7.60 | < 0.001* | $T$ = -5.53 | < 0.001* |
| BMI-SDS at adm./dis. | -2.9 ± 1.2 (-5 – -1.4) | -1.0 ± 0.4 (-1.8 – -0.5) | | $T$ = -11.42 | < 0.001* | $T$ = -8.45 | < 0.001* | $T$ = -5.03 | < 0.001* |
| treatment duration (d) | | 119.2 ± 46.8 (69 – 242) | | | | | | | |
| time from adm. to T1/ time from T2 to dis. (d) | 19.6 ± 12.3 (4 – 60) | 8.1 ± 26.1 (-52 – 52) | | | | | | | |
| **Symptom scales** | | | | | | | | | |
| EDE - mean total score | 3.8 ± 1.2 (1.6 – 5.7) | 2.0 ± 1.1 (0.3 – 4.4) | | | | | | $T$ = 8.00 | < 0.001* |
| EDI-2 - total score | 271.5 ± 80.1 (109 – 436) | 252.5 ± 58.9 (106 – 335) | 187.4 ± 31.9 (115 – 240) | $T$ = 4.56 | < 0.001* | $T$ = 4.42 | < 0.001* | $T$ = 0.88 | 0.383 |
| BDI-2 - total score | 18.4 ± 13.6 (0 – 38) | 13.5 ± 14.2 (0 – 51) | 3.1 ± 2.7 (0 – 10) | $U$ = 83.50 | 0.001* | $U$ = 91.00 | 0.004* | $W$ = 81.50 | 0.073 |

**table S1: Comparisons of demographic and clinical characteristics: cohort 1**

* significant group difference, α = 0.05, $p$-value uncorrected. Group comparisons using independent or paired $t$-tests ($T$), Mann-Whitney-$U$-tests ($U$), or Wilcoxon-tests ($W$) as appropriate.
SD = standard deviation; ED = eating disorder; IQ = intelligence quotient; BMI(-SDS) = body mass index (- standard deviation score); adm. = admission; dis. = discharge; treatment duration = time from admission to discharge; EDE = Eating Disorder Examination; EDI-2 = Eating Disorder Inventory 2; BDI-2 = Beck Depression Inventory 2.
Missing data: T1acu: verbal IQ: $N$ = 6, EDE: $N$ = 3, EDI-2: $N$ = 1, BDI-2: $N$ = 3. T2acu: leptin: $N$ = 1, EDE: $N$ = 3, EDI-2: $N$ = 4, BDI-2: $N$ = 3.

| | T3rec (recovery) | HCrec | Statistics | | | |
|---|---|---|---|---|---|---|
| | $N$ = 21 | $N$ = 22 | T3rec vs. HCrec | | T1/T2acu vs. T3rec | |
| | *mean ± SD (min. – max.)* | *mean ± SD (min. – max.)* | *T* or *U* | *p* | *T* | *p* |
| **General** | | | | | | |
| Age (y) | 22.3 ± 3.3 (17.7 – 31.4) | 22.5 ± 3.5 (16.6 – 31.3) | *T* = -0.20 | 0.843 | | |
| verbal IQ | 108.2 ± 9.5 (97 – 124) | 110.9 ± 12.0 (95 – 130) | *U* = 201.50 | 0.639 | | |
| **ED severity** | | | | | | |
| BMI (kg/m$^2$) | 21.8 ± 2.6 (18.6 – 26.6) | 21.9 ± 2.0 (19.1 – 25.5) | *T* = -0.08 | 0.935 | | |
| BMI-SDS | -0.3 ± 0.8 (-1.4 – 0.9) | -0.2 ± 0.7 (-1.2 – 0.9) | *U* = 217.00 | 0.808 | | |
| leptin (ng/ml) | 12.6 ± 7.2 (3.7 – 26) | 11.1 ± 6.4 (2.8 – 25.6) | *T* = 0.69 | 0.495 | | |
| **ED history** | | | | | | |
| age at ED onset (y) | 14.4 ± 1.6 (11.8 – 17.6) | | | | *T* = -0.75 | 0.461 |
| age at first admission (y) | 15.2 ± 1.6 (12 – 18.4) | | | | *T* = -0.03 | 0.976 |
| BMI at first admission (kg/m$^2$) | 15.7 ± 2.1 (11.3 – 21.3) | | | | *T* = -0.73 | 0.469 |
| BMI-SDS at first admission | -2.7 ± 1.5 (-6.2 – -0.2) | | | | *T* = -0.49 | 0.625 |
| ED duration (y) | 2.1 ± 1.7 (0.1 – 7.1) | | | | | |
| recovery duration (y) | 5.3 ± 3.0 (1.5 – 13.2) | | | | | |
| **Symptom scales** | | | | | | |
| EDE - mean total score | 1.0 ± 1.0 (0.1 – 3.7) | | | | | |
| EDI-2 - total score | 259.8 ± 73.2 (175 – 399) | 194.6 ± 32.8 (151 – 263) | *U* = 91.50 | 0.002* | | |
| BDI-2 - total score | 10.7 ± 11.5 (0 – 37) | 3.9 ± 3.9 (0 – 12) | *U* = 131.50 | 0.041* | | |

**table S2: Comparisons of demographic and clinical characteristics: cohort 2**

* significant group difference, α = 0.05, *p*-value uncorrected. Group comparisons using independent *t*-tests (*T*) or Mann-Whitney-*U*-tests (*U*), as appropriate.
SD = standard deviation; ED = eating disorder; IQ = intelligence quotient; BMI(-SDS) = body mass index (- standard deviation score); adm. = admission; dis. = discharge; ED duration = time from symptom onset to last discharge (if symptom onset not available, from first admission); recovery duration = time from last inpatient discharge or last underweight state to examination; EDE = Eating Disorder Examination; EDI-2 = Eating Disorder Inventory 2; BDI-2 = Beck Depression Inventory 2.
Missing data: T3rec: age at ED onset: *N* = 12, BMI-SDS at first admission: *N* = 1. HCrec: verbal IQ: *N* = 1, EDI-2: *N* = 2, BDI-2: *N* = 1.

**table S3: Additional clinical characteristic of the study sample**

EDE = Eating Disorder Inventory, AN = Anorexia nervosa. AN characteristics as recorded by the EDE were rated as present if reported for at least 1 day of the month prior to the interview. Missing data: T1/T2acu: EDE: $N$ = 3. HCacu: menstruation: $N$ = 1. T3rec: number of past inpatient admissions: $N$ = 1, comorbidities: $N$ = 3, menstruation: $N$ = 2.

| | **Cohort 1** | | | **Cohort 2** | |
|---|---|---|---|---|---|
| | **T1acu** | **T2acu** | **HCacu** | **T3rec** | **HCrec** |
| | *N (%)* | *N (%)* | *N (%)* | *N (%)* | *N (%)* |
| **Number of past inpatient admissions** | | | | | |
| 0 | 19 (86.4) | 0 | 22 (100) | 0 | 22 (100) |
| 1 | 3 (13.6) | 18 (85.7) | 0 | 10 (47.6) | 0 |
| 2 | 0 | 3 (14.3) | 0 | 6 (28.6) | 0 |
| ≥ 3 | 0 | 0 | 0 | 4 (19) | 0 |
| **EDE: AN characteristics** | | | | | |
| bingeing | 1 (4.5) | 1 (4.8) | *n.a.* | 0 | *n.a.* |
| self-induced vomiting | 1 (4.5) | 1 (4.8) | *n.a.* | 0 | *n.a.* |
| use of laxatives | 1 (4.5) | 0 | *n.a.* | 0 | *n.a.* |
| excessive exercising | 14 (63.6) | 8 (38.1) | *n.a.* | 5 (23.8) | *n.a.* |
| **Comorbidities** | | | | | |
| Major depression | 1 (4.5) | | 0 | 3 (14.3) | 0 |
| Minor depression | 15 (68.2) | | 0 | 0 | 0 |
| OCD | 2 (9.1) | | 0 | 0 | 0 |
| Anxiety disorders | 3 (13.6) | | 0 | 5 (23.8) | 0 |
| Adjustment disorder | 1 (4.5) | | 0 | 0 | 0 |
| No comorbidity | 3 (13.6) | | 22 (100) | 12 (57.1) | 22 (100) |
| **Medication** | | | | | |
| antidepressants | 0 | 5 (23.8) | 0 | 3 (14.3) | 0 |
| antipsychotics | 4 (18.2) | 4 (19) | 0 | 0 | 0 |
| no medication | 18 (81.8) | 15 (71.4) | 22 (100) | 18 (85.7) | 22 (100) |
| **Menstruation** | | | | | |
| primary amenorrhoea | 6 (27.3) | 6 (28.6) | 1 (4.5) | 0 | 0 |
| secondary amenorrhoea | 15 (68.2) | 12 (57.1) | 0 | 0 | 0 |
| menstruation | 1 (4.5) | 3 (13.6) | 20 (90.9) | 19 (90.5) | 22 (100) |

| | T1acu vs. HCacu | | T2acu vs. HCacu | | T3rec vs. HCrec | | T1acu vs. T2acu | |
|---|---|---|---|---|---|---|---|---|
| | *T* or *U* | *p* | *T* or *U* | *p* | *T* or *U* | *p* | *T* or *W* | *p* |
| **FWD** | | | | | | | | |
| translation | *U* = 174.00 | 0.114 | *U* = 177.00 | 0.196 | *U* = 231.00 | 1.000 | *T* = 0.04 | 0.969 |
| rotation | *T* = 0.38 | 0.708 | *T* = 0.63 | 0.535 | *T* = -0.91 | 0.368 | *T* = -0.50 | 0.619 |
| **Signal intensities** | | | | | | | | |
| grey matter | *T* = -2.28 | 0.028* | *T* = -1.72 | 0.093 | *T* = 0.25 | 0.801 | *T* = -0.50 | 0.625 |
| white matter | *U* = 148.00 | 0.027* | *T* = -1.16 | 0.252 | *T* = 0.38 | 0.705 | *W* = 68.00 | 0.103 |
| cerebrospinal fluid | *T* = -0.62 | 0.537 | *T* = 0.21 | 0.832 | *T* = 0.87 | 0.388 | *T* = -1.02 | 0.320 |

**table S4: Comparison of frame-wise displacement and global signal intensities**

* significant group difference, α = 0.05, p-value uncorrected. Comparisons using independent or paired t-tests (T), Mann-Whitney-U-tests (U) or Wilcoxon-tests (W), as appropriate.
Frame-wise displacement (FWD) was averaged from 3 translation and 3 rotation parameters. Maximum FWD for each group: translation: T1acu: 1.14 mm, T2acu: 1.29 mm, HCacu: 2.68 mm, T3rec: 0.65 mm, HCrec: 0.44 mm. rotation: T1acu: 0.03°, T2acu: 0.03°, HCacu: 0.17°, T3rec: 0.01°, HCrec: 0.01°. Signal intensities calculated from realigned and normalized functional images using tissue type masks thresholded at a probability of > 0.8.

| Contrast | NBS | | Seed-to-ROI | | | | |
|---|---|---|---|---|---|---|---|
| | Size | $p^{1}$ | seed ROI | degree change | distant ROIs | $T$ | $p^{2}$ |
| **T1acu vs. HCacu** | 460 | 0.022 | LH_Default_PFC_4 (Frontal_Sup_2_L) | 28 | RH_DorsAttn_Post_1 (Temporal_Mid_R) | -4.08 | < 0.001 |
| | | | | | RH_Vis_6 (Calcarine_R) | -3.91 | < 0.001 |
| **T1acu vs. T2acu** | 596 | 0.009 | LH_SomMot_2 (Insula_L) | 29 | LH_Default_PFC_6 (Frontal_Mid_2_L) | -5.58 | < 0.001 |
| | | | | | LH_Limbic_OFC_1 (OFC_Med_L) | -4.60 | < 0.001 |
| | | | | | RH_Cont_PFCl_3 (Frontal_Mid_2_R) | -4.53 | < 0.001 |

### table S5: Network Based Statistics

[1] α = 0.05, *p*-value family wise error corrected [2] α = 0.05/115, *p*-value uncorrected.
Significant subnetworks resulting from the NBS procedure. Reported are network-sizes (number of connections) and associated *p*-values estimated by permutation testing. Also reported are the regions of interest (ROIs) with the strongest decrease in node degree and functional connections to other regions that differed significantly between groups. Independent comparisons included age as covariate. Regions of Interest were derived from the Schaefer et al. (2018) and the Neuromophometrics, Inc atlases. ROIs from the Automated Anatomic Labelling atlas (Rolls et al., 2020) corresponding to the centroid coordinates of the Schaefer et al.-atlas are reported in brackets.

| Original contrast | Cluster or network name | T1acu vs. HCacu | | | T2acu vs. HCacu | | | T3rec vs. HCrec | | | T1acu vs. T2acu | | |
|---|---|---|---|---|---|---|---|---|---|---|---|---|---|
| | | *F* | *p* | *d* | *F* | *p* | *d* | *F* | *p* | *d* | *T* | *p* | *d* |
| **Network Based Statistics NBS** | | | | | | | | | | | | | |
| **T1acu vs. HCacu** | T1acu < HCacu | ° | ° | -2.17 | 8.29 | 0.006* | -0.90 | 0.66 | 0.420 | *n.s.* | -5.19 | < 0.001* | -1.13 |
| | T1acu > HCacu | ° | ° | 1.19 | 8.44 | 0.006* | 0.89 | 1.65 | 0.207 | *n.s.* | 1.00 | 0.329 | *n.s.* |
| **T1acu vs. T2acu** | T1acu < T2acu | 13.00 | < 0.001* | -1.16 | 2.63 | 0.113 | *n.s.* | 0.00 | 0.949 | *n.s.* | ° | ° | -1.73 |
| | T1acu > T2acu | 0.08 | 0.783 | *n.s.* | 31.84 | < 0.001* | -1.73 | 1.48 | 0.230 | *n.s.* | ° | ° | 1.75 |
| **Overlap T1acu < HCacu/ T2acu** | | 35.90 | < 0.001* | -1.90 | 0.58 | 0.451 | *n.s.* | 0.41 | 0.528 | *n.s.* | -8.12 | < 0.001* | -1.77 |
| **Global Correlation GC** | | | | | | | | | | | | | |
| **T1acu < HCacu** | prefrontal GC | ° | ° | -1.47 | 3.66 | 0.063 | *n.s.* | 0.07 | 0.795 | *n.s.* | -2.09 | 0.050 | *n.s.* |
| | sensorimotor GC[a] | ° | ° | -1.21 | 0.31 | 0.580 | *n.s.* | 1.04 | 0.313 | *n.s.* | -3.10 | 0.006* | -0.68 |
| **T1acu < T2acu** | sensorimotor GC[b] | 7.79 | 0.008* | -0.91 | 0.39 | 0.538 | *n.s.* | 1.00 | 0.325 | *n.s.* | ° | ° | -0.87 |
| | insular GC | 6.10 | 0.018 | *n.s.* | 1.13 | 0.294 | *n.s.* | 0.43 | 0.513 | *n.s.* | ° | ° | -0.88 |
| **Overlap T1acu < HCacu/ T2acu** | | 13.13 | < 0.001* | -1.16 | 0.00 | 0.991 | *n.s.* | 0.93 | 0.340 | *n.s.* | -3.97 | < 0.001* | -0.87 |
| **Integrated Local Correlation LC** | | | | | | | | | | | | | |
| **T1acu < HCacu** | sensorimotor LC[a] | ° | ° | -1.49 | 2.32 | 0.136 | *n.s.* | 0.16 | 0.688 | *n.s.* | -3.89 | < 0.001* | -0.85 |
| | prefrontal LC | ° | ° | -1.71 | 17.7 | < 0.001* | -1.13 | 1.42 | 0.240 | *n.s.* | -2.02 | 0.057 | *n.s.* |
| | precuneal LC | ° | ° | -1.65 | 5.82 | 0.021 | *n.s.* | 0.25 | 0.618 | *n.s.* | -2.53 | 0.020 | *n.s.* |
| **T1acu < T2acu** | sensorimotor LC[b] | 11.70 | 0.001* | -1.06 | 0.09 | 0.770 | *n.s.* | 0.36 | 0.552 | *n.s.* | ° | ° | -1.34 |
| | temporal LC | 5.51 | 0.024 | *n.s.* | 3.83 | 0.057 | *n.s.* | 0.05 | 0.822 | *n.s.* | ° | ° | -1.54 |
| | fusiform LC | 5.70 | 0.022 | *n.s.* | 5.70 | 0.022 | *n.s.* | 0.02 | 0.890 | *n.s.* | ° | ° | -1.31 |
| **T1acu x T3rec** | (frontal LC) | 18.39 | < 0.001* | -1.65 | 1.80 | 0.187 | *n.s.* | 3.60 | 0.065 | *n.s.* | -4.38 | < 0.001* | -0.96 |
| **Overlap T1acu < HCacu/ T2acu** | | 19.42 | < 0.001* | -1.38 | 0.38 | 0.542 | *n.s.* | 0.18 | 0.674 | *n.s.* | -5.29 | < 0.001* | -1.15 |
| **fractional Amplitude of Low Frequency Fluctuations fALFF** | | | | | | | | | | | | | |
| **T1acu < HCacu** | parietal fALFF | ° | ° | -1.99 | 8.87 | 0.005* | -0.94 | 1.29 | 0.262 | *n.s.* | -3.07 | 0.006* | -0.67 |
| | calcarine fALFF | ° | ° | -1.53 | 1.70 | 0.199 | *n.s.* | 0.76 | 0.390 | *n.s.* | -2.74 | 0.013 | *n.s.* |
| **T1acu < T2acu** | precuneal fALFF | 11.51 | 0.002* | -1.07 | 0.79 | 0.378 | *n.s.* | 0.39 | 0.534 | *n.s.* | ° | ° | -1.17 |
| | temporal fALFF | 8.02 | 0.007* | -0.80 | 3.58 | 0.066 | *n.s.* | 0.71 | 0.404 | *n.s.* | ° | ° | -1.57 |
| **Overlap T1acu < HCacu/ T2acu** | | 21.61 | < 0.001* | -1.40 | 0.06 | 0.810 | *n.s.* | 0.38 | 0.542 | *n.s.* | -5.12 | < 0.001* | -1.12 |

**table S6: Post-hoc comparisons of resting-state group differences**

* significant group difference, α = 0.0125 (Bonferroni-corrected per modality and contrast), *p*-value uncorrected. ° Group comparisons that would have been circular and therefore were not reported.
Results of post-hoc comparisons of primary analyses. For NBS, functional connectivity of each connection included in significant NBS-subnetworks was averaged for the whole network, separately for positive and negative connections. For voxel-wise measures, values of every voxel included in significant clusters were averaged per cluster. The networks or clusters stem from the contrasts listed on the left side; averaged values were compared between all other groups to understand temporal development of resting-state properties that were decreased in acute patients. To assess whether functional connections or clusters that were decreased in both contrasts showed a similar pattern of normalization, overlapping negative connections or voxels were identified, averaged and compared as stated above. Independent group comparisons using analyses of covariances (age was included as covariate), T1acu vs. T2acu comparisons using paired *t*-tests.
*d* = Cohen's *d* (Hedge's *g* corrected). Superscript a and b refer to the corresponding clusters stemming from T1acu < HCacu and T1acu < T2acu contrast, respectively.

| **Contrast** | **NBS** | | **Seed-to-ROI** | | | | |
|---|---|---|---|---|---|---|---|
| | Size | $p^1$ | seed ROI | degree change | distant ROIs | $T$ | $p^2$ |
| **T1acu vs. HCacu** | 500 | 0.020 | LH_Default_PFC_4 (Frontal_Sup_2_L) | 30 | RH_DorsAttn_Post_1 (Temporal_Mid_R) | -4.19 | 0.018 |
| | | | | | RH_Vis_6 (Calcarine_R) | -4.10 | 0.023 |
| | | | | | LH_Vis_6 (Calcarine_L) | -4.00 | 0.032 |

**table S7: Network-Based Statistics controlled for frame-wise displacement**

[1] α = 0.05, *p*-value family wise error corrected. [2] α = 0.05/115, *p*-value uncorrected.
Significant subnetworks resulting from the NBS procedure while including two frame-wise displacement (FWD) covariates (averaged translation and averaged rotation) additionally to age. See table S5.

| Original contrast | Cluster or network name | T1acu vs. HCacu | | T2acu vs. HCacu | |
|---|---|---|---|---|---|
| | | *F* | *p* | *F* | *p* |
| **Network Based Statistics NBS** | | | | | |
| **T1acu vs. HCacu** | T1acu < HCacu | 15.20 | 0.001* | 9.23 | 0.007* |
| | T1acu > HCacu | 2.74 | 0.115 | 2.79 | 0.113 |
| **Global Correlation GC** | | | | | |
| **T1acu < HCacu** | prefrontal GC | 14.26 | 0.001* | | |
| | sensorimotor GC | 3.58 | 0.075 | | |
| **Integrated Local Correlation LC** | | | | | |
| **T1acu < HCacu** | sensorimotor LC | 6.67 | 0.019* | | |
| | prefrontal LC | 16.15 | < 0.001* | 6.49 | 0.021* |
| | precuneal LC | 13.71 | 0.002* | | |
| **T1acu x T3rec** | (frontal LC) | 5.60 | 0.029* | | |
| **fractional Amplitude of Low Frequency Fluctutations** | | | | | |
| **T1acu < HCacu** | parietal fALFF | 8.28 | 0.010* | 3.77 | 0.069 |
| | calcarine fALFF | 5.31 | 0.033* | | |

**table S8: Post-hoc comparisons of resting-state group differences controlled for verbal IQ**

* significant group difference, α = 0.05, *p*-value uncorrected.

Results of post-hoc comparisons of resting-state results, while including verbal intelligence quotient (IQ) as covariate in addition to age. IQ was available for only 16 out of 22 patients with acute AN. Only comparisons showing significant differences in resting-state measures were repeated. See table S6.

| Original contrast | Cluster or network name | T1acu vs. T2acu | |
|---|---|---|---|
| | | *F* | *p* |
| **Network Based Statistics NBS** | | | |
| **T1acu vs. T2acu** | T1acu < T2acu | 15.00 | 0.001* |
| | T1acu > T2acu | 22.72 | < 0.001* |
| **Global Correlation GC** | | | |
| **T1acu < T2acu** | sensorimotor GC[b] | 4.05 | 0.059 |
| | insular GC | 3.11 | 0.094 |
| **Integrated Local Correlation LC** | | | |
| **T1acu < T2acu** | sensorimotor LC[b] | 4.56 | 0.046* |
| | temporal LC | 4.35 | 0.051 |
| | fusiform LC | 5.41 | 0.031* |
| **fALFF** | | | |
| **T1acu < T2acu** | precuneal fALFF | 7.70 | 0.012* |
| | temporal fALFF | 7.56 | 0.013* |

**table S9: Post-hoc comparisons of resting-state group differences controlled for admission-scan-delay**

* significant group difference, α = 0.05, *p*-value uncorrected. Results of post-hoc comparisons of resting-state results, while including admission-scan-delay (time between inpatient admission and actual T1 scan) as covariate in addition to age in repeated measurement analyses of covariances. Only comparisons showing significant differences in resting-state measures were repeated. See table S6.

| Original contrast | Cluster or network name | T1acu vs. HCacu | | T2acu vs. HCacu | | T1acu vs. T2acu | |
|---|---|---|---|---|---|---|---|
| | | *F* | *p* | *F* | *p* | *F* | *p* |
| **Network Based Statistics NBS** | | | | | | | |
| **T1acu vs. HCacu** | T1acu < HCacu | 16.67 | < 0.001* | 7.03 | 0.012* | 7,39 | 0,014 |
| | T1acu > HCacu | 2.63 | 0.113 | 0.70 | 0.410 | | |
| **T1acu vs. T2acu** | T1acu < T2acu | 4.20 | 0.047* | | | 20.30 | < 0.001* |
| | T1acu > T2acu | | | 12.80 | < 0.001* | 21.81 | < 0.001* |
| **Global Correlation GC** | | | | | | | |
| **T1acu < HCacu** | prefrontal GC | 8.98 | 0.005* | | | | |
| | sensorimotor GC[a] | 2.18 | 0.147 | | | 1.12 | 0.303 |
| **T1acu < T2acu** | sensorimotor GC[b] | 1.16 | 0.289 | | | 2.09 | 0.165 |
| | insular GC | | | | | 6.37 | 0.021* |
| **Integrated Local Correlation LC** | | | | | | | |
| **T1acu < HCacu** | sensorimotor LC[a] | 2.64 | 0.112 | | | 5,04 | 0.037* |
| | prefrontal LC | 11.50 | 0.002* | 7.09 | 0.011* | | |
| | precuneal LC | 0.94 | 0.338 | | | | |
| **T1acu < T2acu** | sensorimotor LC[b] | 1.58 | 0.216 | | | 14.83 | 0.001* |
| | temporal LC | | | | | 16.24 | < 0.001* |
| | fusiform LC | | | | | 8.99 | 0.007* |
| **T1acu x T3rec** | (frontal LC) | 4.49 | 0.040* | | | 8.43 | 0.009* |
| **fractional Amplitude of Low Frequency Fluctuations fALFF** | | | | | | | |
| **T1acu < HCacu** | parietal fALFF | 8.59 | 0.006* | 5.13 | 0.029* | 1,52 | 0.233 |
| | calcarine fALFF | 2.41 | 0.128 | | | | |
| **T1acu < T2acu** | precuneal fALFF | 2.00 | 0.165 | | | 15.12 | < 0.001* |
| | temporal fALFF | 0.27 | 0.607 | | | 15.70 | < 0.001* |

**table S10: Post-hoc comparisons of resting-state group differences controlled for BMI-SDS**

***** significant group difference, α = 0.05, *p*-value uncorrected.
Results of post-hoc comparisons of resting-state results significantly differing between groups while including the body mass index - standard deviation score (BMI-SDS) as covariate in addition to age in (repeated measures) analyses of covariances. Only comparisons showing significant differences in resting-state measures were repeated. See table S6.

| Original contrast | Cluster or network name | T1acu vs. HCacu | | | T2acu vs. HCacu | | | T1acu vs. T2acu | | |
|---|---|---|---|---|---|---|---|---|---|---|
| | | *F* | *p* | *d* | *F* | *p* | *d* | *T* | *p* | *d* |
| **Network Based Statistics NBS** | | | | | | | | | | |
| **T1acu vs. HCacu** | T1acu < HCacu | 47.16 | < 0.001* | -2.24 | 6.46 | 0.016* | -0.89 | -4.17 | 0.001* | -1.16 |
| | T1acu > HCacu | 12.50 | 0.001* | 1.16 | 14.31 | < 0.001* | 1.24 | | | |
| **T1acu vs. T2acu** | T1acu < T2acu | 15.10 | < 0.001* | -1.33 | | | | -7.01 | < 0.001* | -1.94 |
| | T1acu > T2acu | | | | 34.87 | < 0.001* | -1.96 | 5.52 | < 0.001* | 1.53 |
| **Global Correlation GC** | | | | | | | | | | |
| **T1acu < HCacu** | prefrontal GC | 18.01 | < 0.001* | -1.42 | | | | | | |
| | sensorimotor GC[a] | 14.86 | < 0.001* | -1.30 | | | | -3.42 | 0.005* | -0.95 |
| **T1acu < T2acu** | sensorimotor GC[b] | 9.24 | 0.004* | -1.05 | | | | -4.41 | < 0.001* | -1.22 |
| | insular GC | | | | | | | -3.67 | 0.003* | -1.02 |
| **Integrated Local Correlation LC** | | | | | | | | | | |
| **T1acu < HCacu** | sensorimotor LC[a] | 20.43 | < 0.001* | -1.50 | | | | -4.02 | 0.002* | -1.11 |
| | prefrontal LC | 29.46 | < 0.001* | -1.70 | 12.14 | 0.001* | -0.99 | | | |
| | precuneal LC | 36.61 | < 0.001* | -1.89 | | | | | | |
| **T1acu < T2acu** | sensorimotor LC[b] | 10.06 | 0.003* | -1.07 | | | | -7.86 | < 0.001* | -2.18 |
| | temporal LC | | | | | | | -5.67 | < 0.001* | -1.57 |
| | fusiform LC | | | | | | | -5.93 | < 0.001* | -1.64 |
| **T1acu x T3rec** | (frontal LC) | 16.11 | < 0.001* | -1.36 | | | | -3.67 | 0.003* | -1.02 |
| **fractional Amplitude of Low Frequency Fluctuations fALFF** | | | | | | | | | | |
| **T1acu < HCacu** | parietal fALFF | 40.29 | < 0.001* | -2.10 | 15.11 | < 0.001* | -1.29 | -3.23 | 0.007* | -0.90 |
| | calcarine fALFF | 28.49 | < 0.001* | -1.78 | | | | | | |
| **T1acu < T2acu** | precuneal fALFF | 14.03 | < 0.001* | | | | | -4.27 | 0.001* | -1.19 |
| | temporal fALFF | 7.97 | 0.008* | | | | | -5.84 | < 0.001* | -1.62 |

**table S11: Post-hoc comparisons of resting-state group differences excluding participants taking psychoactive medication**

* significant group difference, $\alpha = 0.05$, *p*-value uncorrected.
Results of post-hoc comparisons of resting-state results significantly differing between groups while excluding participants taking psychoactive medication. Group-sizes: T1acu: $N = 18$, T2acu: $N = 15$, T1acu vs. T2acu: $N = 13$. No healthy control took psychoactive medication (table S3). Only comparisons showing significant differences in resting-state measures were repeated. See table S6.

| Original contrast | Cluster or network name | T1acu vs. HCacu | | T2acu vs. HCacu | | T1acu vs. T2acu | |
|---|---|---|---|---|---|---|---|
| | | *U* | *p* | *U* | *p* | *W* | *p* |
| **Network Based Statistics NBS** | | | | | | | |
| **T1acu vs. HCacu** | T1acu < HCacu | 23 | < 0.001* | 98 | < 0.001* | 6 | < 0.001* |
| | T1acu > HCacu | 88 | < 0.001* | 123 | 0.008* | | |
| **T1acu vs. T2acu** | T1acu < T2acu | 84 | < 0.001* | | | 0 | < 0.001* |
| | T1acu > T2acu | | | 52 | < 0.001* | 230 | < 0.001* |
| **Global Correlation GC** | | | | | | | |
| **T1acu < HCacu** | prefrontal GC | 56 | < 0.001* | | | | |
| | sensorimotor GC[a] | 75 | < 0.001* | | | 33 | 0.003* |
| **T1acu < T2acu** | sensorimotor GC[b] | 94 | < 0.001* | | | 17 | < 0.001* |
| | insular GC | | | | | 26 | < 0.001* |
| **Integrated Local Correlation LC** | | | | | | | |
| **T1acu < HCacu** | sensorimotor LC[a] | 71 | < 0.001* | | | 20 | < 0.001* |
| | prefrontal LC | 56 | < 0.001* | 106 | 0.002* | | |
| | precuneal LC | 46 | < 0.001* | | | | |
| **T1acu < T2acu** | sensorimotor LC[b] | 113 | 0.002* | | | 3 | < 0.001* |
| | temporal LC | | | | | # | |
| | fusiform LC | | | | | 0 | < 0.001* |
| **T1acu x T3rec** | (frontal LC) | 81 | < 0.001* | | | 15 | < 0.001* |
| **fractional Amplitude of Low Frequency Fluctuations fALFF** | | | | | | | |
| **T1acu < HCacu** | parietal fALFF | 35 | < 0.001* | 114 | 0.004* | 33 | 0.003* |
| | calcarine fALFF | 61 | < 0.001* | | | | |
| **T1acu < T2acu** | precuneal fALFF | 101 | < 0.001* | | | 5 | < 0.001* |
| | temporal fALFF | 147 | 0.025* | | | # | |

**table S12: Post-hoc comparisons of resting-state group differences using non-parametric statistics**

* significant group difference, α = 0.05, p-value uncorrected.
Results of post-hoc comparisons of resting-state results significantly differing between groups while using Mann-Whitney-*U*- or Wilcoxon-tests (*W*) tests to control for the potential influence of data outliers. Outliers were defined as values exceeding 2 standard deviations from the group-wise mean. Except for the comparisons marked by "#", in at least one of the included groups outliers (maximum $N = 2$) were identified. Note that in these analyses we did not control for age because of the statistical tests applied. See table S6.

| Contrast | Cluster name | Permutation statistics | | | T statistics (peak) | | | | AAL regions | |
|---|---|---|---|---|---|---|---|---|---|---|
| | | Threshold | Size | $p$ [1] | $T$ | coordinates | | | peak region | regions covered over 10% |
| **Global Correlation GC** | | | | | | | | | | |
| **T1acu < HCacu** | prefrontal GC | 439 | 1610 | 0.010 | 5.46 | -21 | 66 | -3 | Frontal_Sup_2_L | Frontal_Sup_2_L, Frontal_Sup_2_R, Frontal_Sup_Medial_R, Frontal_Sup_Medial_L, ACC_pre_L, ACC_pre_R, Frontal_Med_Orb_R, Frontal_Med_Orb_L, |
| | sensorimotor GC[a] | | 871 | 0.021 | 4.37 | 12 | -12 | 72 | Supp_Motor_Area_R | Supp_Motor_Area_R, Supp_Motor_Area_L, Paracentral_Lobule_L, Cingulate_Mid_L, Cingulate_Mid_R, Parietal_Sup_R |
| **T1acu < T2acu** | sensorimotor GC[b] | 416 | 913 | 0.019 | 4.91 | 27 | -21 | 72 | Precentral_R | Supp_Motor_Area_L, Paracentral_Lobule_L, Paracentral_Lobule_R, Postcentral_R, Precentral_R, |
| | insular GC | | 507 | 0.039 | 4.65 | 3 | 6 | 0 | Putamen_R | Rolandic_Oper_R, Insula_R, Putamen_R, Pallidum_R |
| **Integrated Local Correlation LC** | | | | | | | | | | |
| **T1acu < HCacu** | sensorimotor LC[a] | 223 | 654 | 0.011 | 5.24 | -24 | -27 | 69 | Precentral_L | Supp_Motor_Area_L, Supp_Motor_Area_R, Paracentral_Lobule_L |
| | prefrontal LC | | 292 | 0.042 | 5.1 | 24 | 60 | 15 | Frontal_Sup_2_R | Frontal_Sup_2_R |
| | precuneal LC | | 289 | 0.043 | 4.77 | 3 | -66 | 21 | Precuneus_R | Cingulate_Post_L |
| **T1acu < T2acu** | sensorimotor LC[b] | 192 | 656 | 0.011 | 6.02 | -6 | -15 | 48 | Supp_Motor_Area_L | Supp_Motor_Area_L, Precentral_L Paracentral_Lobule_L, Cingulate_Mid_L, Supp_Motor_Area_R |
| | temporal LC | | 304 | 0.029 | 6.84 | -45 | -30 | 0 | Temporal_Mid_L | Temporal_Mid_L |
| | fusiform LC | | 285 | 0.031 | 5.47 | 45 | -45 | -24 | Fusiform_R | Fusiform_R, ParaHippocampal_R |
| **T1acu x T3rec** | (frontal LC) | 332 | 380 | 0.046 | 3.97 | 6 | 30 | 42 | Frontal_Sup_Medial_R | Supp_Motor_Area_L, Supp_Motor_Area_R |
| **fractional Amplitude of Low Frequency Fluctuations fALFF** | | | | | | | | | | |
| **T1acu < HCacu** | parietal fALFF | 223 | 377 | 0.022 | 4.76 | -48 | -45 | 36 | Parietal_Inf_L | Parietal_Inf_L, Parietal_Sup_L, Angular_L |
| | calcarine fALFF | | 310 | 0.029 | 4.09 | -3 | -72 | 18 | Calcarine_L | Calcarine_L, Cingulate_Post_L |
| **T1acu < T2acu** | precuneal fALFF | 203 | 327 | 0.028 | 6.3 | -3 | -60 | 15 | Precuneus_L | Precuneus_L, Cingulate_Post_L |
| | temporal fALFF | | 210 | 0.048 | 5.63 | -48 | -33 | 0 | Temporal_Mid_L | Temporal_Mid_L |

**table S13: Voxel-wise resting-state measures**

[1] $\alpha = 0.05$, $p$-value family-wise error corrected.
The statistic was based on a combination of an uncorrected voxel-wise threshold of $p < 0.01$ and an exact permutation based (1000 permutations) cluster threshold of $p < 0.05$. Reported are cluster thresholds, sizes of the resulting clusters and associated $p$-values. Additionally, we listed $T$-value, coordinates and corresponding regions in the Automated Anatomic Labelling atlas (AAL, Rolls et al, 2020) of each cluster's peak coordinates. Independent comparisons included age as covariate.

| Contrast | Corresponding cluster name | Permutation statistics | | | T statistics (peak) | | | | AAL regions | |
|---|---|---|---|---|---|---|---|---|---|---|
| | | Threshold | Size | $p$ [1] | $T$ | coordinates | | | peak region | regions covered over 10% |
| | | | | | **Global Correlation GC** | | | | | |
| **T1acu < HCacu** | prefrontal GC | 446 | 1806 | 0.010 | 5.85 | -21 | 66 | -3 | Frontal_Sup_2_L | Frontal_Sup_2_L, Frontal_Sup_2_R, Frontal_Sup_Medial_R, Frontal_Sup_Medial_L, ACC_pre_L, ACC_pre_R, ACC_sup_L, ACC_sub_l Frontal_Med_Orb_R, Frontal_Med_Orb_L, OFCant_L, |
| | sensorimotor GC[a] | | 510 | 0.045 | 4.26 | -3 | -24 | 75 | Paracentral_Lobule_L | Paracentral_Lobule_L, Cingulate_Mid_L |
| | | | | | **Integrated Local Correlation LC** | | | | | |
| **T1acu < HCacu** | sensorimotor LC[a] | 212 | 415 | 0.021 | 4.22 | 0 | -18 | 48 | Supp_Motor_Area_L | Supp_Motor_Area_L, Supp_Motor_Area_R |
| | prefrontal LC | | 309 | 0.029 | 4.80 | 24 | 60 | 18 | Frontal_Sup_2_R | Frontal_Sup_2_R |
| | precuneal LC | | 299 | 0.030 | 4.41 | 3 | -66 | 21 | Precuneus_R | Cingulate_Post_L |
| | - | | 278 | 0.032 | 4.58 | 42 | 0 | -36 | Temporal_Inf_R | Temporal_Pole_Mid_R, Frontal_Inf_Orb_2_R |
| | - | | 255 | 0.037 | 5.14 | 3 | 60 | -9 | Frontal_Med_Orb_R | Frontal_Med_Orb_L, Frontal_Med_Orb_R, ACC_sub_L |
| | - | | 227 | 0.045 | 4.86 | 39 | -63 | 30 | Angular_R | (Temporal_Mid_R) |
| **T1acu x T3rec** | (frontal LC) | 365 | *n.s.* | | | | | | | |
| | | | | | **fractional Amplitude of Low Frequency Fluctuations fALFF** | | | | | |
| **T1acu < HCacu** | parietal fALFF | 216 | 233 | 0.044 | 4.38 | -54 | -60 | 42 | Angular_L | Parietal_Sup_L, Angular_L |
| | calcarine fALFF | | 264 | 0.039 | 4.31 | -3 | -72 | 18 | Calcarine_L | Calcarine_L, Cingulate_Post_L |

**table S14: Voxel-wise resting-state measures controlled for frame-wise displacement**

[1] $\alpha = 0.05$, $p$-value family-wise error corrected.
Two frame-wise displacement (FWD) covariates (averaged translation and averaged rotation) were, in addition to age, included in the comparisons of independent groups. See table S13.

| Modality | Cluster or network name | BMI-SDS | | leptin | | EDE | | EDI-2 | | BDI-2 | | untreated[1] | | T1-T2-time[2] | |
|---|---|---|---|---|---|---|---|---|---|---|---|---|---|---|---|
| | | *r* | *p* | *r* | *p* | *r* | *p* | *r* | *p* | *r* | *p* | *r* | *p* | *r* | *p* |
| **baseline (T1acu)** | | | | | | | | | | | | | | | |
| **NBS** | T1acu < HCacu | 0.12 | 0.609 | 0.07 | 0.757 | -0.16 | 0.515 | 0.53 | 0.014* | 0.19 | 0.435 | -0.06 | 0.791 | | |
| **NBS** | T1acu > HCacu | -0.05 | 0.832 | -0.13 | 0.563 | 0.19 | 0.430 | 0.23 | 0.306 | 0.34 | 0.149 | 0.11 | 0.617 | | |
| **GC** | prefrontal GC | 0.01 | 0.978 | -0.13 | 0.578 | -0.12 | 0.614 | 0.35 | 0.117 | -0.10 | 0.696 | 0.05 | 0.826 | | |
| **GC** | sensorimotor GC[a] | 0.22 | 0.318 | 0.31 | 0.158 | -0.12 | 0.611 | 0.10 | 0.675 | 0.05 | 0.830 | -0.11 | 0.629 | | |
| **LC** | sensorimotor LC[a] | 0.24 | 0.272 | 0.15 | 0.505 | -0.09 | 0.724 | -0.01 | 0.982 | 0.00 | 0.989 | -0.01 | 0.972 | | |
| **LC** | prefrontal LC | 0.13 | 0.556 | -0.34 | 0.127 | -0.03 | 0.917 | 0.34 | 0.130 | 0.02 | 0.937 | 0.13 | 0.556 | | |
| **LC** | precuneal LC | 0.49 | 0.020* | 0.36 | 0.104 | 0.09 | 0.727 | 0.43 | 0.049* | 0.46 | 0.050* | -0.03 | 0.892 | | |
| **fALFF** | parietal fALFF | 0.19 | 0.395 | -0.16 | 0.490 | -0.08 | 0.733 | -0.15 | 0.509 | -0.04 | 0.865 | -0.01 | 0.956 | | |
| **fALFF** | calcarine fALFF | 0.18 | 0.427 | 0.12 | 0.599 | -0.02 | 0.941 | 0.16 | 0.482 | 0.24 | 0.313 | 0.07 | 0.763 | | |
| **delta (T2-T1)** | | | | | | | | | | | | | | | |
| **NBS** | T1acu < T2acu | 0.09 | 0.706 | 0.00 | 0.983 | 0.21 | 0.411 | -0.13 | 0.624 | -0.05 | 0.846 | | | 0.24 | 0.304 |
| **NBS** | T1acu > T2acu | -0.06 | 0.798 | -0.31 | 0.180 | 0.01 | 0.968 | 0.16 | 0.552 | -0.08 | 0.771 | | | 0.12 | 0.592 |
| **GC** | sensorimotor GC[b] | 0.30 | 0.185 | 0.05 | 0.849 | 0.11 | 0.656 | -0.31 | 0.234 | -0.19 | 0.480 | | | 0.37 | 0.094 |
| **GC** | insular GC | -0.03 | 0.907 | -0.22 | 0.348 | 0.11 | 0.664 | -0.13 | 0.622 | -0.11 | 0.688 | | | 0.26 | 0.261 |
| **LC** | sensorimotor LC[b] | -0.04 | 0.877 | -0.13 | 0.582 | 0.45 | 0.059 | -0.28 | 0.285 | -0.12 | 0.660 | | | 0.30 | 0.192 |
| **LC** | temporal LC | 0.07 | 0.750 | -0.02 | 0.936 | 0.29 | 0.241 | -0.18 | 0.501 | 0.07 | 0.798 | | | 0.34 | 0.129 |
| **LC** | fusiform LC | 0.21 | 0.365 | -0.07 | 0.754 | 0.45 | 0.062 | -0.27 | 0.292 | -0.11 | 0.672 | | | 0.44 | 0.046* |
| **fALFF** | precuneal fALFF | -0.17 | 0.462 | -0.12 | 0.624 | 0.36 | 0.139 | 0.06 | 0.826 | 0.06 | 0.832 | | | 0.07 | 0.779 |
| **fALFF** | temporal fALFF | 0.12 | 0.608 | -0.08 | 0.749 | 0.13 | 0.619 | -0.30 | 0.241 | -0.09 | 0.740 | | | 0.43 | 0.050* |

**table S15: Correlations of resting-state results with clinical variables**

* significant correlation, α = 0.05, *p*-value uncorrected. [1] Time between symptom onset and inpatient admission. [2] Time between T1acu and T2acu scans. Reported are Pearson correlations (*r*) between results from resting-state analyses (network- or cluster-wise averaged) and clinical variables. The correlations were calculated for the T1acu group (baseline) and between changes in measures from T1 to T2 (delta).
BMI-SDS = body mass index (- standard deviation score); EDE = Eating Disorder Examination; EDI-2 = Eating Disorder Inventory 2; BDI-2 = Beck Depression Inventory 2; NBS = Network Based Statistics; GC = Global Correlation; LC = Integrated Local Correlation; fALFF = fractional Amplitude of Low Frequency Fluctuations. Superscript a and b refer to the corresponding clusters stemming from T1acu < HCacu and T1acu < T2acu contrast, respectively.

| Original contrast | Cluster or network name | T1acu vs. HCacu | | | T2acu vs. HCacu | | | T1acu vs. T2acu | | |
|---|---|---|---|---|---|---|---|---|---|---|
| | | *F* | *p* | *d* | *F* | *p* | *d* | *T* | *p* | *d* |
| **Regions included in Network Based Statistics subnetworks** | | | | | | | | | | |
| **T1acu vs. HCacu** | T1acu < HCacu | 13.52 | < 0.001* | -1.17 | 4.23 | 0.046* | -0.66 | -6.06 | < 0.001* | -1.32 |
| | T1acu > HCacu | 18.33 | <0.001* | -1.32 | 5.85 | 0.020* | -0.76 | | | |
| **T1acu vs. T2acu** | T1acu < T2acu | 13.59 | < 0.001* | -1.17 | | | | -5.99 | < 0.001* | -1.31 |
| | T1acu > T2acu | | | | 3.70 | 0.061 | *n.s.* | -5.75 | < 0.001* | -1.25 |
| **Global Correlation clusters** | | | | | | | | | | |
| **T1acu < HCacu** | prefrontal GC | 7.92 | 0.007* | -0.90 | | | | | | |
| | sensorimotor GC[a] | 7.89 | 0.008* | -0.89 | | | | -3.01 | 0.007* | -0,66 |
| **T1acu < T2acu** | sensorimotor GC[b] | 9.02 | 0.005* | -0.96 | | | | -4.18 | < 0.001* | -0.91 |
| | insular GC | | | | | | | -6.38 | < 0.001* | -1.39 |
| **Integrated Local Correlation clusters** | | | | | | | | | | |
| **T1acu < HCacu** | sensorimotor LC[a] | 7.35 | 0.010* | -0.87 | | | | -2,85 | 0.010* | -0,62 |
| | prefrontal LC | 8.85 | 0.005* | -0.98 | 1.93 | 0.172 | *n.s.* | | | |
| | precuneal LC | 19.12 | < 0.001* | -1.40 | | | | | | |
| **T1acu < T2acu** | sensorimotor LC[b] | 10.52 | 0.002* | -1.03 | | | | -3.11 | 0.006* | -0.68 |
| | temporal LC | | | | | | | -5.45 | < 0.001* | -1.19 |
| | fusiform LC | | | | | | | -4.89 | < 0.001* | -1.07 |
| **T1acu x T3rec** | (frontal LC) | 6.64 | 0.014* | 0.82 | | | | -2.81 | 0.011* | -0.61 |
| **fractional Amplitude of Low Frequency Fluctuations clusters** | | | | | | | | | | |
| **T1acu < HCacu** | parietal fALFF | 16.91 | < 0.001* | -1.32 | 5.90 | 0.020* | -0.78 | -3,89 | < 0.001* | -0,85 |
| | calcarine fALFF | 17.83 | < 0.001* | -1.35 | | | | | | |
| **T1acu < T2acu** | precuneal fALFF | 17.73 | < 0.001* | -1.35 | | | | -6.79 | < 0.001* | -1.48 |
| | temporal fALFF | 15.73 | < 0.001* | -1.21 | | | | -4.85 | < 0.001* | -1.06 |

**table S16: Post-hoc comparisons of voxel-wise grey matter volumes**

* significant group differences, α = 0.05, *p*-value uncorrected.
Results of group comparisons of network- or cluster-wise grey matter volume (GMV), estimated from individual voxel-wise GMV-maps. Only comparisons of values showing significant differences in resting-state measures (see table S6) were conducted. For NBS, GMV of all regions included in the resulting subnetworks was averaged, separately for positive and negative connections. For voxel-wise measures, GMV was averaged per cluster. Independent group comparisons using analyses of covariances (age was included as covariate), T1acu vs. T2acu comparisons using paired *t*-tests.
*d* = Cohen's *d* (Hedge's *g* corrected)*;* Superscript a and b refer to the corresponding clusters stemming from T1acu < HCacu and T1acu < T2acu contrast, respectively.

| Original contrast | Cluster or network name | T1acu vs. HCacu | | | T2acu vs. HCacu | | | T1acu vs. T2acu | | |
|---|---|---|---|---|---|---|---|---|---|---|
| | | *F* | *p* | *d* | *F* | *p* | *d* | *F / T* | *p* | *d* |
| | | | | **Network Based Statistics NBS[1]** | | | | | | |
| **T1acu vs. HCacu** | T1acu < HCacu | 29.34 | < 0.001* | -1.61 | 3.77 | 0.059 | *n.s.* | -4.25 | < 0.001* | -0.93 |
| | T1acu > HCacu | 11.55 | 0.002* | 1.03 | 6.82 | 0.013* | 0.80 | | | |
| **T1acu vs. T2acu** | T1acu < T2acu | 7.41 | 0.009* | -0.88 | | | | -7.45 | < 0.001* | -1.62 |
| | T1acu > T2acu | | | | 30.47 | < 0.001* | -1.69 | 8.18 | < 0.001* | 1.78 |
| | | | | **Global Correlation GC[2]** | | | | | | |
| **T1acu < HCacu** | prefrontal GC | 17.19 | < 0.001* | -1,29 | | | | | | |
| | sensorimotor GC[a] | 11.50 | 0.002* | -1.09 | | | | -2.90 | 0.009* | -0.63 |
| **T1acu < T2acu** | sensorimotor GC[b] | 5.97 | 0.019* | -0.81 | | | | -3.81 | 0.001* | -0.83 |
| | insular GC | | | | | | | -3.75 | 0.001* | -0.82 |
| | | | | **Integrated Local Correlation LC[2]** | | | | | | |
| **T1acu < HCacu** | sensorimotor LC[a] | 14.28 | < 0.001* | -1.20 | | | | -3,18 | 0.005* | -0.69 |
| | prefrontal LC | 25.86 | < 0.001* | -1.44 | 15.07 | < 0.001* | -1.03 | | | |
| | precuneal LC | 17.39 | < 0.001* | -1.21 | | | | | | |
| **T1acu < T2acu** | sensorimotor LC[b] | 7.29 | 0.010* | -0.85 | | | | -5.41 | < 0.001* | -1.18 |
| | temporal LC | | | | | | | -6.52 | < 0.001* | -1.42 |
| | fusiform LC | | | | | | | -5.61 | < 0.001* | -1.22 |
| **T1acu x T3rec** | (frontal LC) | 9.97 | 0.003* | -1.01 | | | | -3.31 | 0.004* | -0.72 |
| | | | | **fractional Amplitude of Low Frequency Fluctuations fALFF[2]** | | | | | | |
| **T1acu < HCacu** | parietal fALFF | 37.13 | < 0.001* | -1.85 | 7.29 | 0.010* | -0.86 | -2,84 | 0.010* | -0.62 |
| | calcarine fALFF | 14.79 | < 0.001* | -1.17 | | | | | | |
| **T1acu < T2acu** | precuneal fALFF | 7.07 | 0.011* | -0.84 | | | | -4.94 | < 0.001* | -1.08 |
| | temporal fALFF | 5.23 | 0.027* | -0.63 | | | | -6.80 | < 0.001* | -1.48 |

**table S17: Post-hoc comparisons of resting-state group differences controlled for (voxel-wise) grey matter volume**

* significant group differences, α = 0.05, *p*-value uncorrected.
Resting-state group differences controlled for grey matter volume (GMV). [1] In case of NBS, averaged GMV was regressed out of the averaged network FC separately for each cohort. [2] For voxel-wise resting-state measures (GC, LC, fALFF) GMV was regressed out of the beta-maps of each measure in a voxel-wise manner, values were averaged per cluster and compared between groups. Only comparisons showing significant differences in resting-state measures (see table S6) were repeated. Independent group comparisons using analyses of covariances (age was included as covariate), T1acu vs. T2acu comparisons using paired *t*-tests. Superscript a and b refer to the corresponding clusters stemming from T1acu < HCacu and T1acu < T2acu contrast, respectively.

| Modality | Cluster or network name | GMV | |
|---|---|---|---|
| | | *r* | *p* |
| **baseline (T1acu)** | | | |
| **NBS** | T1acu < HCacu | 0,18 | 0,417 |
| **NBS** | T1acu > HCacu | 0,25 | 0,257 |
| **GC** | prefrontal GC | 0,14 | 0,531 |
| **GC** | sensorimotor GC[a] | 0,06 | 0,795 |
| **LC** | sensorimotor LC[a] | 0,11 | 0,629 |
| **LC** | prefrontal LC | 0,16 | 0,474 |
| **LC** | precuneal LC | 0,26 | 0,246 |
| **fALFF** | parietal fALFF | -0,04 | 0,871 |
| **fALFF** | calcarine fALFF | 0,16 | 0,483 |
| **delta (T2-T1)** | | | |
| **NBS** | T1acu < T2acu | 0.07 | 0.772 |
| **NBS** | T1acu > T2acu | -0,07 | 0,765 |
| **GC** | sensorimotor GC[b] | 0,00 | 0,995 |
| **GC** | insular GC | -0,09 | 0,682 |
| **LC** | sensorimotor LC[b] | 0,01 | 0,956 |
| **LC** | temporal LC | 0,08 | 0,731 |
| **LC** | fusiform LC | 0,33 | 0,148 |
| **fALFF** | precuneal fALFF | 0,02 | 0,942 |
| **fALFF** | temporal fALFF | 0,05 | 0,827 |

**table S18: Correlations of resting-state results with grey matter volumes**

Correlations (Pearson's *r*) between results from resting-state analyses (network- or cluster-wise averaged) and grey matter volumes (GMV). The correlations were calculated for the T1acu group (baseline) and between changes in measures from T1 to T2 (delta).
Superscript a and b refer to the corresponding clusters stemming from T1acu < HCacu and T1acu < T2acu contrast, respectively.

| | Present study (T1acu) | Biezonski et al., 2016 | Boehm et al., 2014/ Ehrlich et al., 2015/ Geisler et al., 2016 | de la Cruz et al., 2021 | Favaro et al., 2012 |
|---|---|---|---|---|---|
| | | Seed-to-voxel | ICA/ NBS/ GT | Seed-to-voxel, NBS | ICA |
| | *N* = 22 | *N* = 28 | *N* = 35 | *N* = 22 | *N* = 29 |
| | *mean ± SD (min. – max.)* | *mean ± SD (min. – max.)* | *mean ± SD (min. – max.)* | *mean ± SD (min. – max.)* | *mean ± SD (min. – max.)* |
| age at ED onset (y) | 13.9 ± 1.9 (9.1 – 17.9) | 14.9 ± 0.4 (*n.a.*) | 13.5 ± 1.7 (*n.a.*) | *n.a.* | 18.2 ± 4.4 (*n.a.*) |
| age at examination (y) | 15.3 ± 1.9 (10.2 – 18.6) | 19.4 ± 0.4 (*n.a.*) | 16.1 ± 2.6 (12 – 23) | 23.8 ± 7.2 (*n.a.*) | 25.8 ± 6.9 (*n.a.*) |
| BMI at examination (kg/m$^2$) | 15.7 ± 1.5 (13.1 – 18.3) | 17.2 ± 0.3 (*n.a.*) | 14.8 ± 1.3 (*n.a.*) | 15.1 ± 1.4 (*n.a.*) | 14.5 ± 2.3 (*n.a.*) |
| ED duration (y) | 1.6 ± 1.3 (0.5 – 4.8) | 4.5 ± 0.6 (*n.a.*) | 2.6* | *n.a.* | 6.2 ± 6.9 (*n.a.*) |

| | Gaudio et al., 2015[1] | Gaudio et al., 2018[1] | Haynos et al., 2019 | Kullmann et al., 2014 |
|---|---|---|---|---|
| | ICA | NBS | Seed-to-voxel | Degree centrality |
| | *N* = 16 | *N* = 15 | *N* = 19 | *N* = 12 |
| age at ED onset (y) | 15.4 ± 1.6 (*n.a.*) | 15.2 ± 1.6 (*n.a.*) | 14.3* | *n.a.* |
| age at examination (y) | 15.8 ± 1.7 (13 – 18) | 15.7 ± 1.7 (13 – 18) | 22.3 ± 3.9 (*n.a.*) | 23.3 ± 4.7 (*n.a.*) |
| BMI at examination (kg/m$^2$) | 16.2 ± 1.2 (*n.a.*) | 16.1 ± 1.2 (*n.a.*) | 17.0 ± 1.4 (*n.a.*) | 15.5 ± 1.5 (*n.a.*) |
| ED duration (y) | 0.3 ± 0.2 (*n.a.*) | 0.3 ± 0.2 (*n.a.*) | 8.0 ± 3.7 (n.a.) | *n.a.* |

| | Lee et al., 2014 | Phillipou et al., 2016 | Scaife et al., 2017 | Seidel et al., 2019 |
|---|---|---|---|---|
| | Seed-to-voxel | ROI-to-ROI | ICA | fALFF, ReHo |
| | *N* = 18 | *N* = 26 | *N* = 12 | *N* = 74 |
| age at ED onset (y) | 21.4* | 16.0 ± 3.4 (*n.a.*) | 20.1 ± 5.9 (*n.a.*) | 14.5* |
| age at examination (y) | 25.2 ± 4.2 (20 – 35) | 22.8 ± 6.7 (*n.a.*) | 29.4 ± 6.0 (*n.a.*) | 16.0 ± 2.9 (12.1 – 28.5) |
| BMI at examination (kg/m$^2$) | 16.0 ± 1.7 (*n.a.*) | 16.6 ± 1.2 (*n.a.*) | 15.4 ± 1.9 (*n.a.*) | 14.6 ± 1.3 (*n.a.*) |
| ED duration (y) | 3.8 ± 2.6 (*n.a.*) | 6.4 ± 7.4 (*n.a.*) | 10.3 ± 5.2 (*n.a.*) | 1.5 ± 1.9 (*n.a.*) |

**table S19: Sample data from selected current resting-state studies in acute Anorexia nervosa**

Sample data as reported by the authors. [1] participants were drawn from the same sample. * ED duration or age of onset were not reported by the authors, but calculated from age at onset and examination, respectively age at examination and ED duration.
SD = standard deviation; ED = eating disorder; BMI = body mass index; ICA = Independent Component Analysis; NBS = Network Based Statistics; GT = graph theory; fALFF = fractional Amplitude of Low Frequency Fluctuations; ReHo = Regional Homogeneity; *n.a.* = not reported by the authors.

| | Present study (T1/T2acu) | Cha et al., 2016 | Uniacke et al., 2019 |
|---|---|---|---|
| | | Seed-to-voxel | Seed-to-voxel |
| | *N* = 22/21 | *N* = 22 | *N* = 25 |
| | *mean ± SD (min. – max.)* | *mean ± SD (min. – max.)* | *mean ± SD (min. – max.)* |
| age at ED onset (y) | 13.9 ± 1.9 (9.1 – 17.9) | *n.a.* | 15.5* |
| age at first scan (y) | 15.3 ± 1.9 (10.2 – 18.6) | 19.5 ± 2.4 (16 – 25) | 19.1 ± 3.5 (14 – 26) |
| BMI at first scan (kg/m$^2$) | 15.7 ± 1.5 (13.1 – 18.3) | 17.3 ± 1.2 (14.8 – 19.0) | 16.5 ± 2.0 (*n.a.*) |
| time between scans (d) | 90.8 ± 40.8 (41 – 183) | 47.6 ± 10.1 (*n.a.*) | 57.1 ± 21.2 (*n.a.*) |
| BMI at second scan (kg/m$^2$) | 18.2 ± 1.2 (15.0 – 20.1) | 20.0 ± 1.6 (*n.a.*) | 20.8 ± 1.1 (*n.a.*) |
| ED duration (at first scan; y) | 1.6 ± 1.3 (0.5 – 4.8) | *n.a.* | 3.6 ± 2.8 (*n.a.*) |

**table S20: Sample data from current longitudinal resting-state studies in acute and short-term-recovered Anorexia nervosa**

Sample data as reported by the authors. * ED onset age was not directly reported but calculated from age at first scan and ED duration.
SD = standard deviation; ED = eating disorder; BMI = body mass index; *n.a.* = not reported by the authors.

| | Present study (T3rec) | Boehm et al., 2016 | Cowdrey et al., 2014 | Favaro et al., 2012 |
|---|---|---|---|---|
| | | ICA | ICA | ICA |
| | *N* = 21 | *N* = 31 | *N* = 16 | *N* = 16 |
| | *mean ± SD (min. – max.)* | *mean ± SD (min. – max.)* | *mean ± SD (min. – max.)* | *mean ± SD (min. – max.)* |
| inclusion criteria (m) | 12 | 6 | 12 | 6 |
| age at ED onset (y) | 14.4 ± 1.6 (11.8 – 17.6) | 14.4 ± 1.9 (*n.a.*) | 14.7 ± 1.7 (*n.a.*) | 17.9 ± 2.8 (*n.a.*) |
| age at examination (y) | 22.3 ± 3.3 (17.7 – 31.4) | 22.3 ± 3.1 (*n.a.*) | 23.1 ± 3.6 (*n.a.*) | 23.8 ± 4.8 (*n.a.*) |
| ED duration (y) | 2.0 ± 1.7 (0.1 – 7.1) | 3.7 ± 2.7 (*n.a.*) | 3.5 ± 2.4 (*n.a.*) | 2.3 ± 1.7 (*n.a.*) |
| recovery duration (y) | 5.3 ± 3 (1.5 – 13.2) | 4.4 ± 2.8 (0.8 – *n.a.*) | 4.9* | 2.8 ± 2.6 (0.5 – 7.5) |

| | Scaife et al., 2017 | Seidel et al., 2020 | Geisler et al., 2019 |
|---|---|---|---|
| | ICA | fALFF, ReHo, MSSD, DC, VHMC | NBS |
| | *N* = 14 | *N* = 65 | *N* = 55 |
| inclusion criteria (m) | 12 | 6 | 6 |
| age at ED onset (y) | 16.5 ± 2.1 (*n.a.*) | *n.a.* | *n.a.* |
| age at examination (y) | 27 ± 6.5 (*n.a.*) | 22.1 ± 3.4 (15.5 – 29.7) | 22.4 ± 3.3 (*n.a.*) |
| ED duration (y) | 5.8 ± 4.2 (*n.a.*) | *n.a.* | *n.a.* |
| recovery duration (y) | 4.7* | 4.3 ± 3.3 (0.75 – 14) | *n.a.* |

**table S21: Sample data from current resting-state studies in long-term-recovered Anorexia nervosa**

Sample data as reported by the authors. "Inclusion criteria" refers to the minimum recovery time allowed for Anorexia nervosa patients to be included in the studies. *Recovery durations were not directly reported, but calculated from onset age, ED duration and age at examination. All listed studies found alterations in recovered Anorexia nervosa patients compared to healthy controls. However, results were inconsistent and different approaches were used in the single studies. We did not refer to Geisler et al. (2019) in our main text, due to missing methodological overlap.

SD = standard deviation; ED = eating disorder; ICA = Independent Component Analysis; fALFF = fractional Amplitude of Low Frequency Fluctuations; ReHo = Regional Homogeneity; MSSD = Mean-Square Successive Difference; VHMC = Voxel-Mirrored Homotopic Connectivity; DC = Degree Centrality; NBS = Network Based Statistics; *n.a.* = not reported by the authors.

### Supplementary Figures

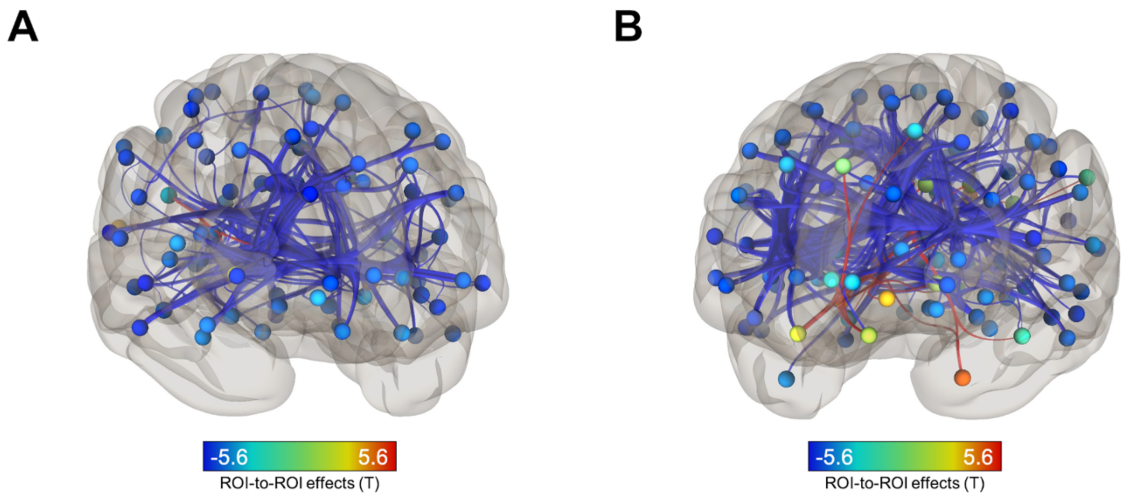


**figure S1: Animated visualisation of Network Based Statistics results**

A: Subnetwork resulting from T1acu vs. HCacu comparison with primarily decreased connections in patients at T1. B: Subnetwork resulting from T1acu vs. T2acu comparison with mainly decreased connections in T1acu. Animated versions of these figures in GIF file format are available separately.

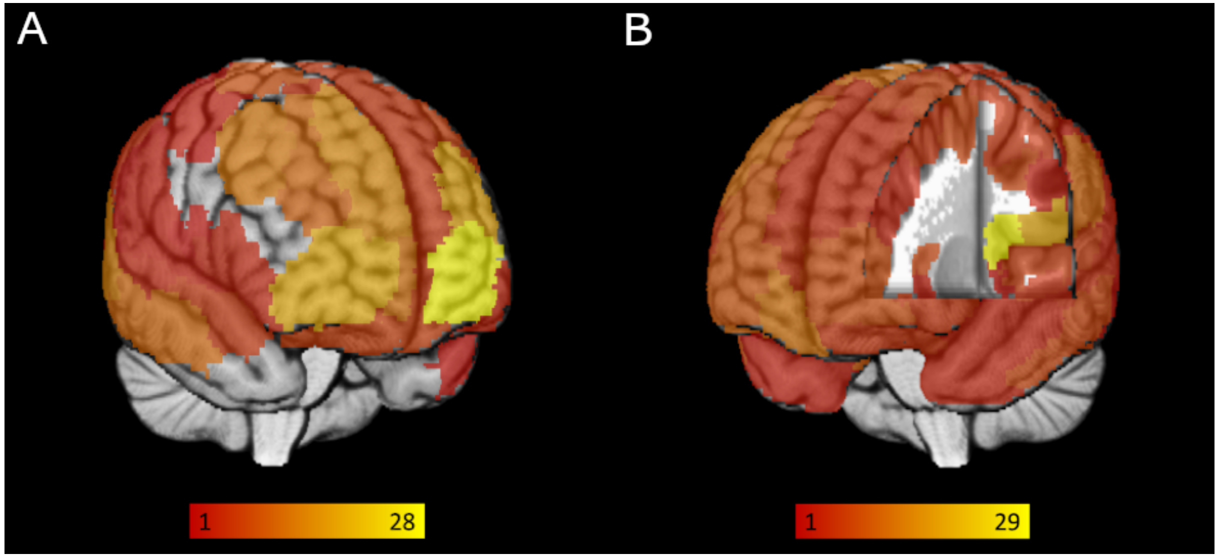


**figure S2: Visualisation of node degree changes in Network Based Statistics subnetworks**

Colour of brain regions reflects extend of changes (decreases) of the degree of each region (yellow = strong decrease) in AN. Node degree refers to the number of connections between a certain node and all other nodes. A: Results from T1acu vs. HCacu comparison. The strongest change was observed in prefrontal regions. B: Results from T1acu vs. T2acu comparison. The insula displayed the largest degree decrease. Animated versions of these figures in GIF file format are available separately.

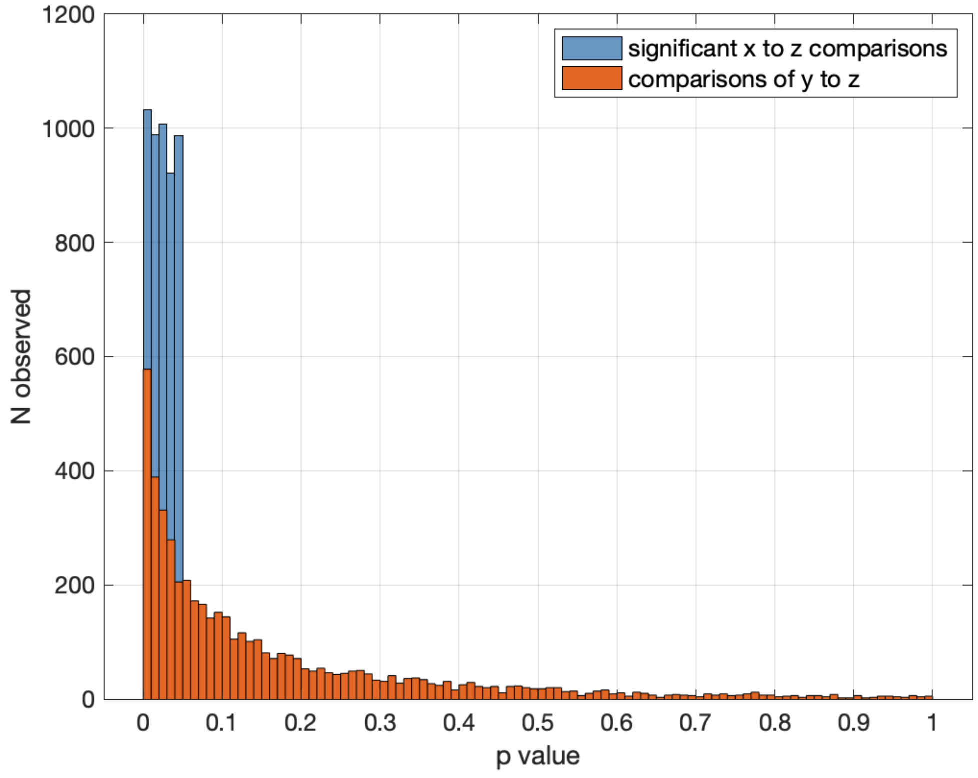


**figure S3: Simulation experiment to assess potential statistical bias affecting post-hoc group comparisons**

Results of a simulation (100.000 repetitions) showing the distribution of *p* values resulting from independent *t*-tests comparing two random vectors y and z (orange bars) if a *t*-test comparing two random vectors x and z returned a *p* value $< 0.05$ (blue bars). Vector lengths resembled group sizes in our study, vectors x and y were generated correlated to each other.

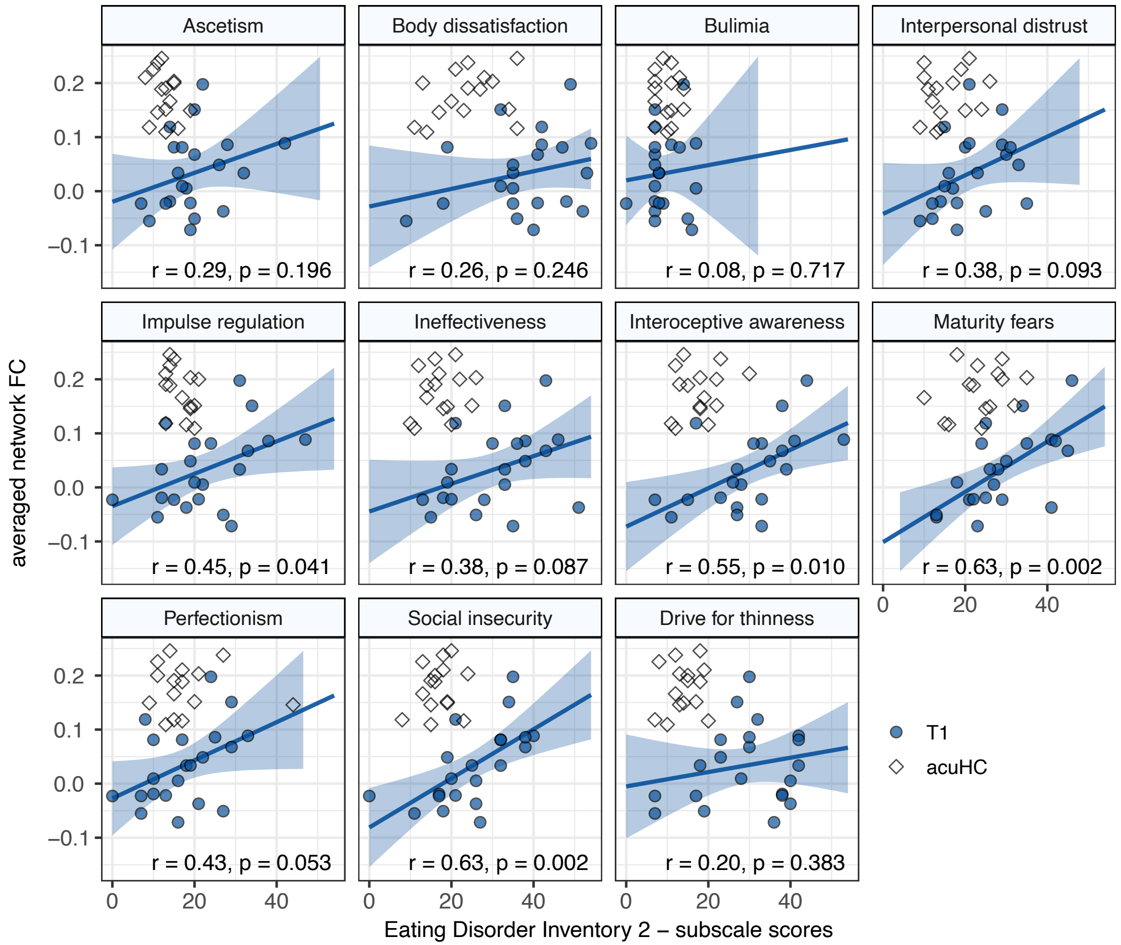


**figure S4: Correlations between T1acu < HCacu network FC and EDI-2 subscales**

Pearson correlations between T1acu < HCacu network functional connectivity (FC) and Eating Disorder Inventory 2 (EDI-2) subscales in acute Anorexia nervosa patients (T1acu). Blue scatter points represent patients with acute AN at T1. Blue lines display the fitted linear regression function, blue areas the corresponding 95% confidence interval. For descriptive purposes, HCacu subjects are shown as white squares, but do not influence the correlation calculation. r = Pearson's *r*.
